# A Genome-wide Association Study of Alzheimer’s Disease and Dementia in a Large Multi-ancestry Military Cohort Identifies Many New Dementia-Associated Loci

**DOI:** 10.64898/2026.05.01.26352216

**Authors:** Richard Sherva, Henry Bayly, Rui Zhang, Kelly Harrington, Jesse Mez, Mark W. Miller, Debby Tsuang, Erika Wolf, Qing Zeng, Yann Le Guen, Marlene Tejeda, the VA Million Veteran Program, the MVP Cognitive Decline, Dementia During Aging Working Group, John Michael Gaziano, Matthew S. Panizzon, Richard L. Hauger, Victoria C. Merritt, Lindsay A. Farrer, Mark W. Logue

## Abstract

**Introduction:** Biobank-scale cohorts of individuals with genetic data and diagnoses of Alzheimer’s disease and related dementias (ADRD) have facilitated the discovery of additional risk loci via meta-analysis, with existing cohorts assembled specifically for ADRD genetic discovery. Cross-ancestry meta-analyses have further elucidated the overall genetic architecture of these dementias. Here, we include for the first time the European ancestry (EA) and Hispanic ancestry (HA) subset of the VA Million Veterans Program (MVP) along with the African ancestry (AA) MVP participants in a meta-analysis with a large-scale EA and AA meta-analysis.

**Methods:** Independent genome-wide association studies (GWASs) were conducted in MVP participants using four phenotypes derived from electronic medical records and surveys: ADRD, prescriptions for common dementia medications, and self-reported maternal and paternal history of dementia (dementia by proxy). These GWASs were repeated in the EA, AA, and HA cohorts. MVP ancestry-specific and cross-ancestry meta-analyses were conducted. These were then meta-analyzed with existing GWAS results. Functionality of the peak variants was explored using brain-derived gene expression data and co-localization analysis.

**Results:** Apart from the *APOE* region, 17, 4, and 3 genome-wide significant (GWS) loci were observed in the MVP EA, AA, and HA meta-analyses, respectively. When we meta-analyzed these with consortium results, we observed 72 loci in the EA GWAS, and 62 lead loci in the cross-ancestry meta-analysis. While most of these loci were known, 27 genes/regions were identified containing variants surpassing genome-wide significance for the first time: 7 EA specific, 12 in the cross-ancestry meta-analysis, and 8 driven by AA and HA cohorts. Several of these are members of pathways containing established ADRD risk genes, and several of the peak SNPs showed evidence for eQTL effects on their respective genes. Several of the novel SNPs showed significant eQTL effects in brain-derived mRNA-seq experiments. Additionally, there was a significant differential expression of the novel gene *PAX7* in ADRD cases and controls.

**Discussion:** MVP represents a large and unique primarily male cohort comprised of US Veterans from a range of backgrounds with a unique set of environmental exposures. The results generated here demonstrate the utility of biobank level cohorts for AD genetic discovery. Furthermore, our discovery of ADRD genes was enhanced by the inclusion of MVP data that provided an increase of underrepresented ancestry groups in contrast to prior cross ancestry GWASs. The new AD risk loci identified present potential new targets for dementia treatment confirmed that future large-scale analyses of AD genetic risk and prediction will be enhanced by the inclusion of MVP data.

## Introduction

Risk of late-onset Alzheimer’s disease (AD) is determined by a complex interaction of health, life history, and genetic risk factors^1,2^. The largest genetic-effect risk locus is the Apolipoprotein E (*APOE*) E4 variant for late-onset AD^3^. Although many other loci have been described in genome-wide association studies (GWASs; e.g. ^4,5^), they have a much smaller individual effect. Although these loci for the most part are not clinically actionable, they can be useful in aggregate^6,7^ and provide important insights into the diverse mechanisms that cause AD pathology^2^. Recent availability of large-scale biobank cohorts with genetic data (e.g. ^8,9^), collaborative consortiums of AD researchers, and proxy dementia information for younger individuals based on reported dementia in parents^10,11^ have boosted the number of individuals included in AD genetic analyses and led to the discovery of nearly 100 AD genetic risk loci in European ancestry (EA) meta-analyses. For example, Bellenguez et al.^4^ identified 74 loci that surpass the genome-wide significance (GWS) threshold of 5.0x10^-8^ in a sample of over 111,000 cases that included both diagnosed and proxy individuals. A subsequent EA meta-analysis included 72,721 AD cases, 55,960 proxy ADRD cases, 614,267 controls and 235,566 proxy controls from 52 studies and identified 91 GWS loci^12^. These meta-analyses are sufficiently powered to detect variants with odds ratios (OR) as small as 1.05 that account for a 5% increase or decrease in risk. These studies have also identified mechanisms including dysregulation of brain immune function, protein degradation and clearance, lipid binding, endosome/lysosome function, Tau phosphorylation in microglia and neurons that contribute to AD pathogenesis.

Recent efforts have focused on increasing sample sizes for non-EA AD study cohorts and incorporating them into ancestry group-specific analyses and cross-ancestry meta-analyses^13^. Expanding the availability of non-EA populations for genetic analysis that are smaller is a critical goal for enhancing AD genetic analyses and discovery. This is important for several reasons. First, there is a notable imbalance between the number of EA individuals included in GWAS studies and all other continental-ancestry groups^13^. This has important ramifications as the effect sizes and the most relevant risk variants in a particular genomic region can differ by ancestry^14^. The performance of genetic risk scores is considerably limited when the population of the GWAS forming the basis of these scores differs in ancestry from the population in which the scores are evaluated^15-17^. Furthermore, due to the small sample sizes, incorporating non-EA GWAS results into risk scores in non-EA populations does not improve their accuracy^18^. The accuracy of these risk scores can be boosted when using a weighted sum polygenic risk score (PRS) information to combine genetic risk scores across ancestry^19^.

Fewer than ten well-characterized and robustly replicated AD risk loci have been identified in African ancestry (AA) cohorts^20^, the second largest ancestry group included in AD GWASs. Although the sample sizes of large-scale genetic analysis of Hispanic ancestry (HA)^21^, East Asian ancestry^22-25^, and other ancestry cohorts are several-fold smaller than those of EA and AA cohorts, they have revealed some novel causal variants in and near previously established AD loci as well as novel AD loci^26^. Cross-ancestry genetic analyses of AD have also been conducted, and advances in methodology for combining data from genetically diverse populations have improved power for discovery^27,28^. A recent cross-ancestry GWAS included 109,479 ADRD cases, 74,141 proxy cases, 2,631,507 controls, and identified 118 GWS loci in a multi-ancestry analysis and 9 additional loci in ancestry-specific analyses^29^. Nonetheless, small sample sizes of non-EA cohorts can be enlarged, many more novel AD loci and variants are likely to be detected.

In this study, we leveraged data from The US Veterans Affairs (VA) Office of Research and Development Million Veteran Program (MVP), which is one of the world’s largest, diverse genetic-data repositories, to boost power to detect genetic associations for AD and dementia. MVP is a biobank-scale study with more than one million United States military Veteran participants at present. Nearly half of the MVP cohort is age 65 or older and hence at increased risk for AD and other forms of dementia. MVP demographics roughly mirror Veterans receiving VA services as a whole; while MVP participants are primarily EA males, MVP also includes a substantial proportion of AA and HA participants. Importantly for this study, the MVP AA and HA cohorts are among the largest available from a single source. In previous work, we were able to discover novel variants in our AA dementia GWAS due to the MVP AA sample size being double that for the previously largest AA dementia study^20^. Here, we present results from ancestry-specific and multi-ancestry ADRD GWAS that includes the largest number of dementia and proxy dementia cases (n=205,550) and largest number of non-EA cases (n=19,867).

## Results

Table 1 shows the demographic characteristics of the MVP sample by harmonized ancestry and race/ethnicity (HARE^30^) ancestry and sex. As a cohort of veterans with previous military service, it is heavily weighted towards males. Across the three ancestry groups, the MVP sample included 30,460 ADRD cases, 7,886 AD medication cases, and 53,055 proxy dementia cases, of which 63% were maternal. Across all meta-analyses, 27 loci surpassed genome wide significance for the first time. Regional Manhattan plots for these regions are shown in supplementary materials.

**Table 1.**
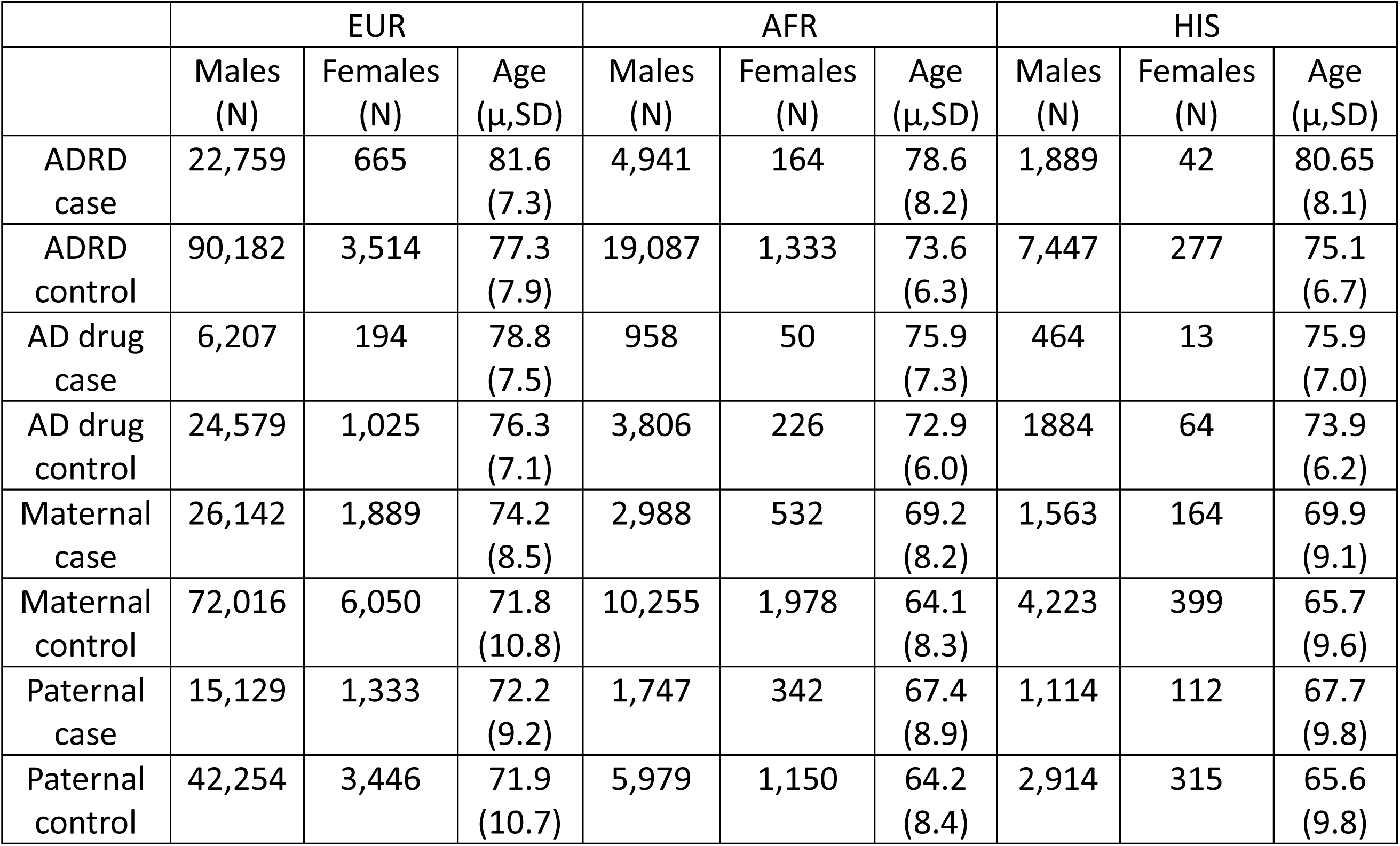
Demographic characteristics of the Million Veteran Program cohort.

### Associations for ADRD in the European ancestry cohorts

In the combined MVP EA cohort (International Classification of Diseases (ICD) and medication diagnosed ADRD and the maternal and paternal proxy AD cohorts), we identified 17 peak variants (including previously reported loci, Figure 1, Table S1). Genomic inflation was modest (λ =1.143), and LD-score regression intercept of 1.094 indicates that this inflation is due to the polygenic effect of variants. We identified GWS associations with ADRD in the MVP EA dataset prior to meta-analysis with existing samples (Table 2) in 5 genes: Chimerin 2 (*CHN2*; rs2040755, p=4.15x10^-8^), Importin 7 (*IPO7*; rs11603265, p=9.3x10^-9^), Galanin And GMAP Prepropeptide (*GAL*; rs34802137, p=2.90x10^-9^), Retinoblastoma 1 (*RB1*; rs117081003, p=1.25x10^-8^), and Myeloid Associated Differentiation Marker Like 2 (*PYCR1*:*MYADML2*; rs564421236, p=1.40x10^-10^).

**Figure 1.**
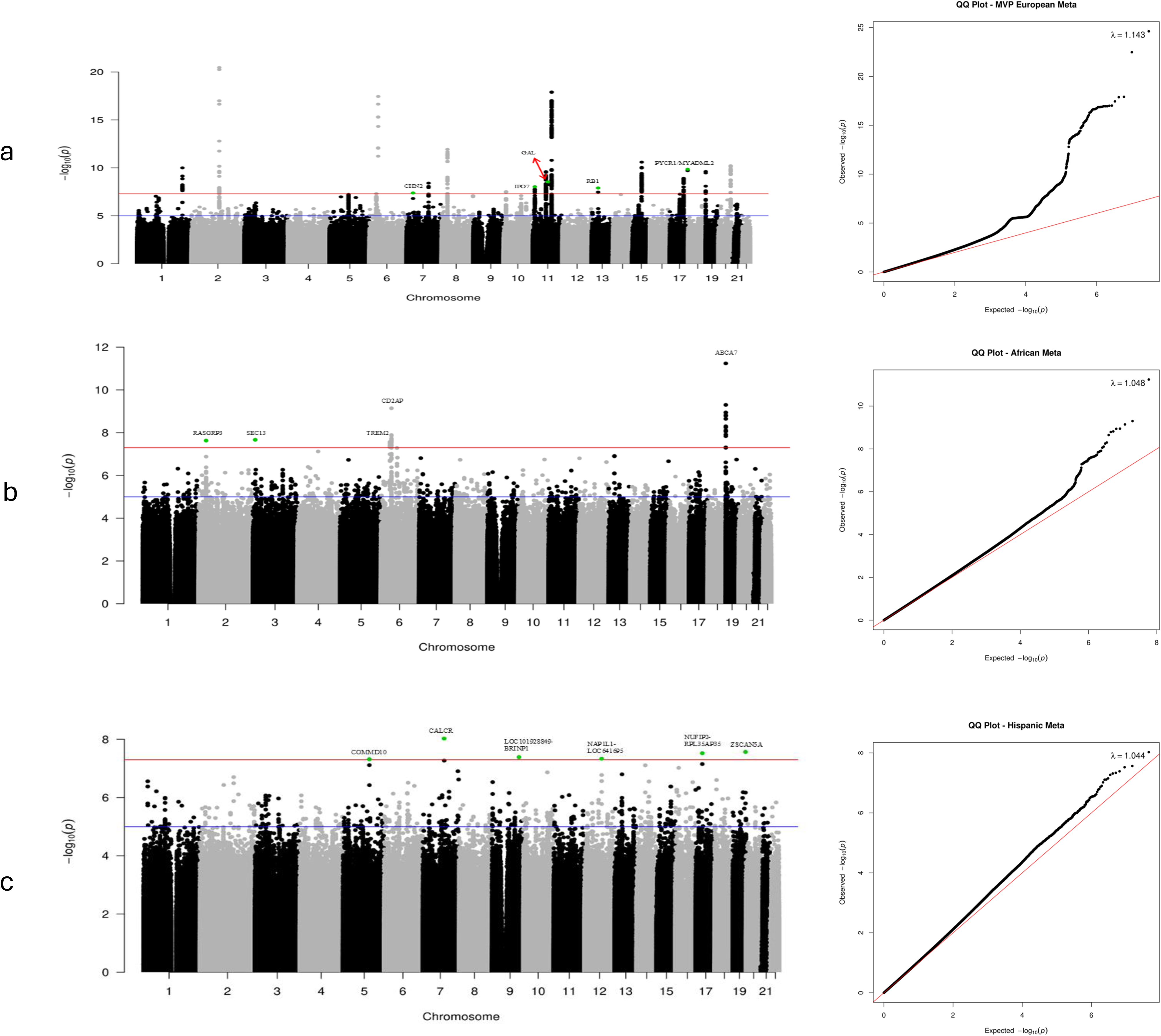
Chromosomal position is on the X-axis and -log10 p-values are on the Y-axis. The red line indicates the significance threshold of p=5.0x10^-8^. The green points indicate SNPs that were not previously genome wide significant (p<5.0x10^-8^).

**Table 2.** Novel genome-wide significant risk loci in the meta-analysis of MVP European ancestry participants.

| Chr | BP | SNP | Gene | EA | NEA | EA Freq | P ADRD | P ADMED | P MAT | P PAT | OR Meta | P Meta | Dir |
| --- | --- | --- | --- | --- | --- | --- | --- | --- | --- | --- | --- | --- | --- |
| 7 | 29384314 | rs2040755 | CHN2 | T | C | 0.013 | 2.24E-05 | 8.04E-02 | 1.08E-01 | 4.12E-03 | 0.83 | 4.15E-08 | ---- |
| 11 | 9363835 | rs11603265 | IPO7 | A | G | 0.43 | 5.61E-07 | 1.52E-01 | 1.63E-01 | 9.04E-03 | 0.96 | 9.30E-09 | ---- |
| 11 | 68694513 | rs34802137 | GAL | T | C | 0.08 | 2.59E-04 | 1.45E-02 | 4.80E-04 | 2.09E-02 | 0.92 | 2.90E-09 | ---- |
| 13 | 48347156 | rs117081003 | RB1 | T | C | 0.008 | 9.65E-03 | 1.56E-03 | 1.09E-03 | 1.15E-03 | 0.77 | 1.25E-08 | ---- |
| 17 | 81942223 | rs564421236 | PYCR1/MYADML2 | A | G | 0.006 | 1.92E-06 | 5.24E-05 | 1.11E-01 | 8.07E-02 | 1.37 | 1.40E-10 | ++++ |

EA=effect allele
NEA=non-effect allele
EA freq=frequency of the effect allele
P ADRD=p-value from the ADRD GWAS
P ADMED=p-value from the AD drug GWAS
P MAT=p-value from the maternal proxy GWAS
P PAT=p-value from the paternal proxy GWAS
OR meta=odds ratio from the meta-analysis
P Meta=p-value from the meta-analysis

After meta-analysis of results from the MVP datasets and the large EA GWAS conducted by Bellenguez et al, 10,018 GWS SNPs across 72 independent genomic loci (including previously reported, Figure 2, Table S2) were identified. Genomic inflation was modest (λ =1.118), and LD-score regression intercept of 1.099 indicated that the inflation was due to the polygenic effect of variants. The following 13 novel GWS regions were identified (Table 3): Arginine-Glutamic Acid Dipeptide Repeats (*RERE*; rs159963, p=1.25x10^-8^), Transmembrane Protein 163 (*TMEM163*; rs35564151, p=6.09x10^-9^), Islet Cell Autoantigen 1 Like (*ICA1L*; rs139738276, p=1.45x10^-9^), Tetratricopeptide Repeat And Ankyrin Repeat Containing 1 (*TRANK1*; rs11129735, p=3.37x10^-9^), Calpastatin (*CAST*, rs5866663, p=2.38x10^-8^), TNFAIP3 Interacting Protein 1 (*TNIP1*; rs871269, p=4.32x10^-10^), Ubiquitin Domain Containing 1 (*UBTD1*; rs11592144, p=2.56x10^-9^), Zinc Finger Protein 143 (*ZNF143*; rs2290425, p=1.94x10^-8^), Serine Protease 23 (*PRSS23*; rs12799759, p=3.57x10^-8^), FA Complementation Group A (*FANCA*; rs17226232, p=1.17x10^-8^), LLGL Scribble Cell Polarity Complex Component 1 (*LLGL1*; rs12603466, p=1.18x10^-8^), Spindle And Kinetochore Associated Complex Subunit 2 (*SKA2*; rs7214098, p=9.52x10^-9^), and Zinc Fingers And Homeoboxes 3 (*ZHX3*; rs17265513, p=1.43x10^-8^).

**Figure 2.**
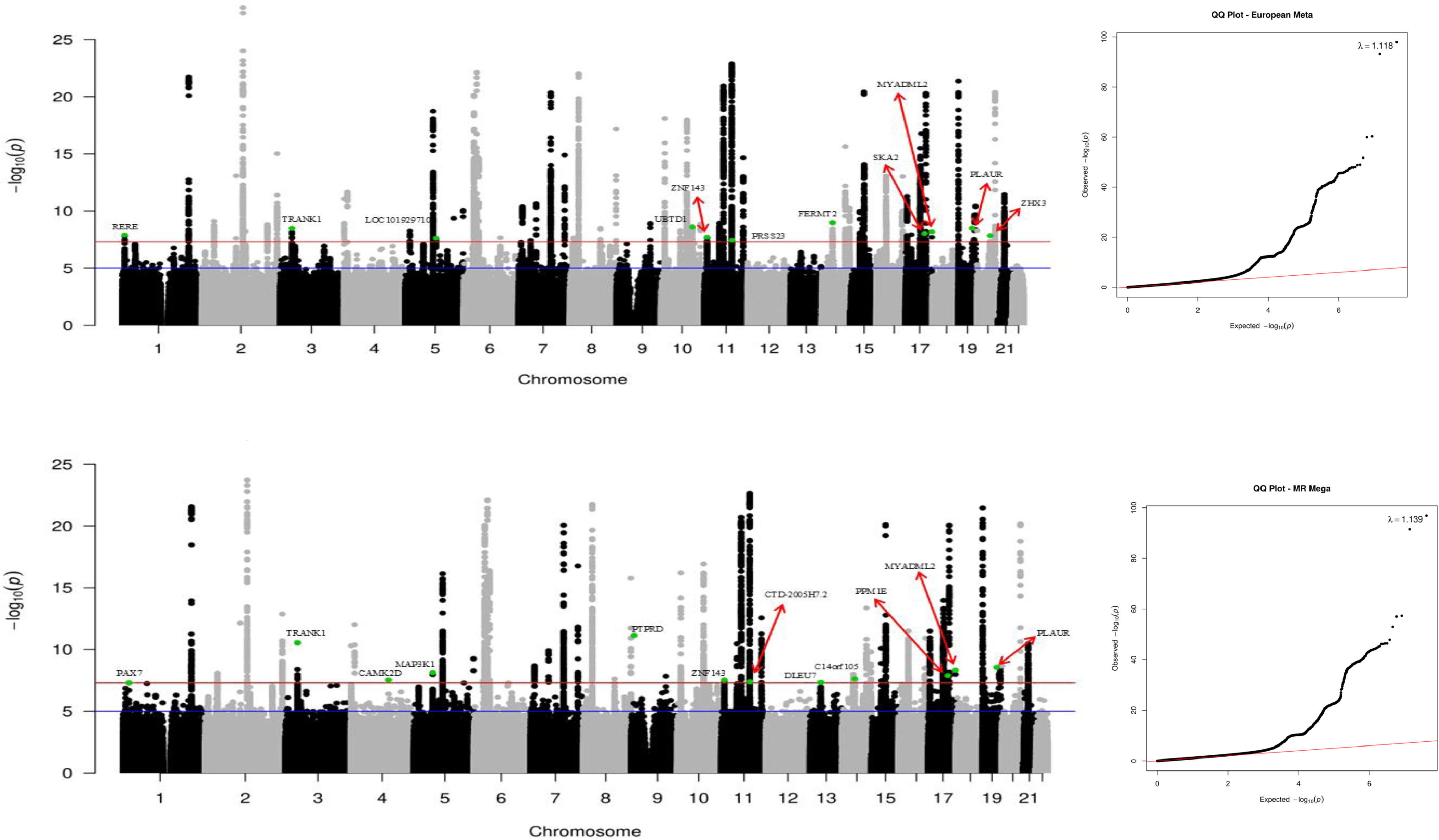
Chromosomal position is on the X-axis and -log10 p-values are on the Y-axis. The red line indicates the significance threshold of p=5.0x10^-8^. The green points indicate SNPs that were not previously genome wide significant (p<5.0x10^-8^).

**Table 3.**
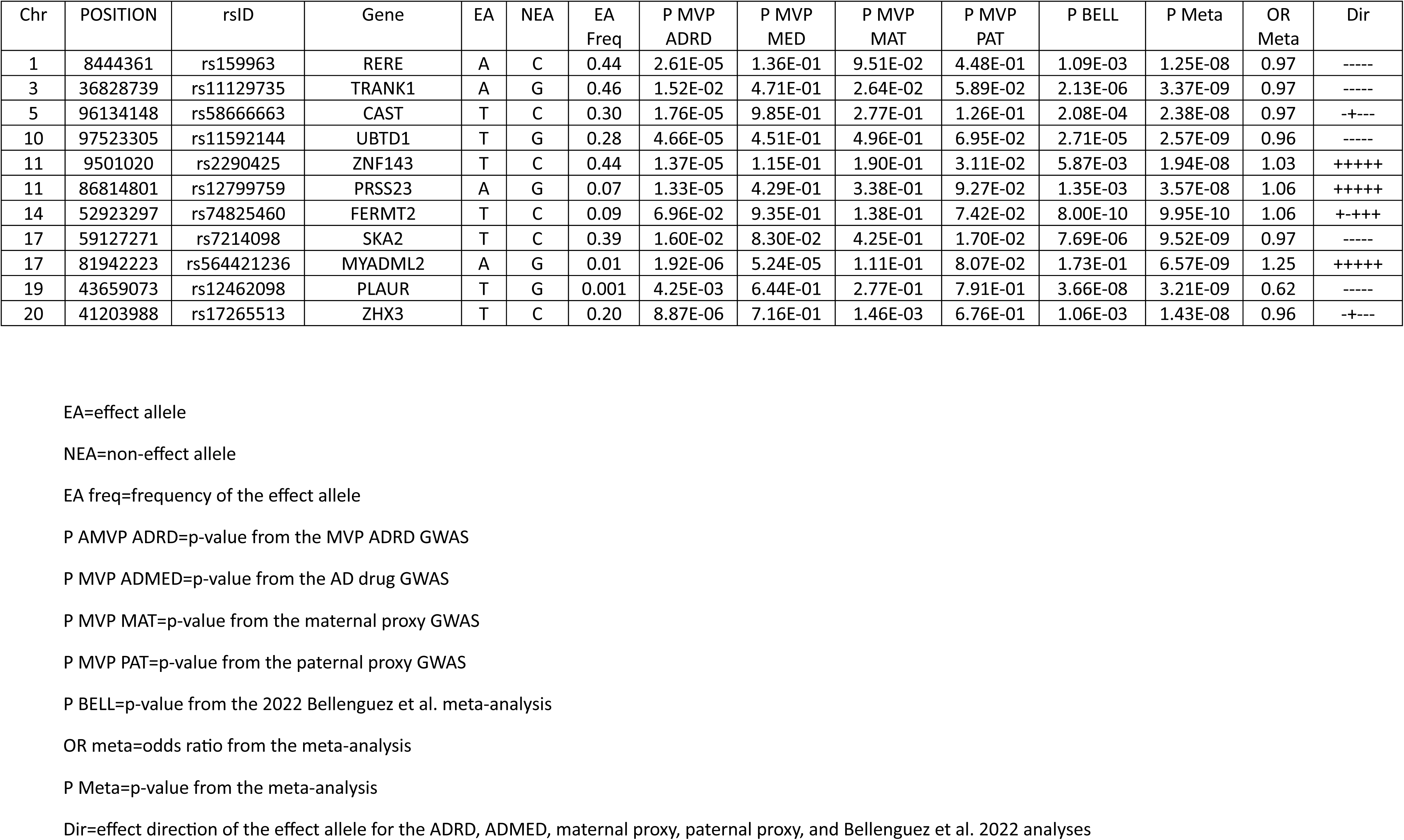
Novel genome-wide significant risk loci in the meta-analysis of European Ancestry participants from MVP and Bellenguez et al. 2022.

### Associations for ADRD in the AA and HA ancestry groups

In the combined AA cohorts from the MVP and Alzheimer’s Disease Genetics Consortium (ADGC), there was modest evidence of genomic inflation (λ=1.048, Figure 1). GWS associations were observed with 120 SNPs in 4 independent regions, including SNPs at 2 novel loci, Ras Guanine Nucleotide Releasing Protein 3 (*RASGRP3*; rs13415828, MAF=0.14, OR=0.87, p=2.34x10^-8^) and SEC13 homolog, nuclear pore and COPII component (*SEC13*; rs376570302, MAF=0.005, OR=0.31, p=2.14x10^-8^). The *RASGRP3* finding was supported by both the MVP (p=4.45x10^-6^) and ADGC datasets (p=0.0009) and was not identified in our previous AA meta-analysis^20^, possibly due to imputation in the MVP dataset using a newer TOPMed reference panel. There was no result for the *SEC13* SNP in ADGC AAs. A total of 403 GWS SNPs in 6 novel loci were identified in the HA MVP participants, with little genomic inflation (λ=1.044, Figure 1, Table 4) GWS. These loci included COMM Domain Containing 10 (*COMMD10*; rs534469689, OR=6.53, p=4.85x10^-8^), Calcitonin Receptor (*CALCR*; rs191185084, OR=5.14, p=9.40x10^-9^), overlapping genes Epididymal Protein 13 (*EDDM13*) and Zinc Finger and SCAN Domain Containing 5A (*ZSCAN5A*), and intergenic regions between *LOC101928849* and *BRINP1* (rs77206068, OR=1.68, p=4.09x10^-8^), 48 kb upstream of *NAP1A1* (rs35823174, OR=2.75, p=4.62x10^-8^) and between *NUFIP2* and *TAOK1* (rs58145907, OR=0.54, p=3.00x10^-8^), and Zinc Finger And SCAN Domain Containing 5A (*ZSCAN5A*; rs529405075, OR=1.65, p=2.72x10^-8^).

**Table 4.** Novel genome-wide significant risk loci in the meta-analysis of MVP Hispanic ancestry participants.

| Chr | POSITION | rsID | Gene | EA | NEA | EA Freq | P ADRD | P MED | P MAT | P PAT | OR Meta | P Meta | Dir |
| --- | --- | --- | --- | --- | --- | --- | --- | --- | --- | --- | --- | --- | --- |
| 5 | 116202400 | rs534469689 | COMMD10 | A | C | 0.001 | 1.22E-03 | 1.98E-01 | 2.47E-01 | 8.80E-07 | 6.53 | 4.85E-08 | ++++ |
| 7 | 93453552 | rs191185084 | CALCR | A | G | 0.002 | 2.99E-04 | 3.50E-01 | 3.64E-02 | 3.62E-07 | 5.14 | 9.40E-09 | ++++ |
| 9 | 119020405 | rs77206068 | LOC101928849-BRINP1 | A | G | 0.04 | 8.18E-08 | 9.96E-02 | 1.26E-01 | 4.69E-01 | 1.68 | 4.09E-08 | ++++ |
| 12 | 76131802 | rs35823174 | NAP1L1-LOC641695 | G | GC | 0.004 | 3.82E-04 | 1.12E-01 | 1.11E-02 | 7.00E-04 | 2.75 | 4.62E-08 | ++++ |
| 17 | 29333743 | rs58145907 | NUFIP2-RPL35AP35 | A | C | 0.01 | 3.51E-06 | 1.91E-01 | 5.35E-02 | 3.22E-02 | 0.54 | 3.00E-08 | ---- |
| 19 | 56278426 | rs529405075 | ZSCAN5A | A | G | 0.02 | 2.90E-06 | 1.77E-01 | 1.27E-01 | 1.15E-02 | 1.65 | 2.72E-08 | ++++ |

EA=effect allele
NEA=non-effect allele
EA freq=frequency of the effect allele
P ADRD=p-value from the ADRD GWAS
P ADMED=p-value from the AD drug GWAS
P MAT=p-value from the maternal proxy GWAS
P PAT=p-value from the paternal proxy GWAS
OR meta=odds ratio from the meta-analysis
P Meta=p-value from the meta-analysis

### Cross-ancestry meta-analyses

In the CA meta-analysis, 7,050 SNPs across 62 regions (including previously reported, Figure 2, Table S3) were GWS. The λ was 1.139. An LD-score regression-based measure of inflation was not conducted due to inherent differences in LD patterns across ancestry groups. Eight of the GWS loci identified in the CA meta-analysis were novel (Table 5): *PAX7* (rs11804731, p=4.95x10^-8^), Calcium/Calmodulin Dependent Protein Kinase II Delta (*CAMK2D*; rs111561336, p=2.76x10^-11^), Mitogen-Activated Protein Kinase Kinase Kinase 1 (*MAP3K1*; rs3736430, p=7.58x10^-9^), *OR2E1P* (rs2859374, p=3.42x10^-8^), *LOC102724775*, (rs7110253, p=4.06x10^-8^), Deleted In Lymphocytic Leukemia 7 (*DLEU7*; rs2812243, p=4.55x10^-8^), *C14orf105* (rs268836, p=2.37x10^-8^), Protein Phosphatase Methylesterase 1 (*PPM1E*; rs9893351, p=1.29x10^-8^), and Protein Tyrosine Phosphatase Receptor Type D (*PTPRD*; rs113482775, p=7.24x10^-12^The *PTPRD* finding was driven entirely by the AA and HA groups and the *PAX7* and *C14orf105* findings were driven primarily by HA participants. The signal in *TRANK1* improved from 2.04x10^-8^ in EA to 2.26x10^-11^ after including AA and HA results, although the signal was only observed in AA (p=1.41x10^-6^). The complete results for all novel and previously reported GWS loci across all ancestry groups, phenotypes and cohorts are shown in Table S4.

**Table 5.**
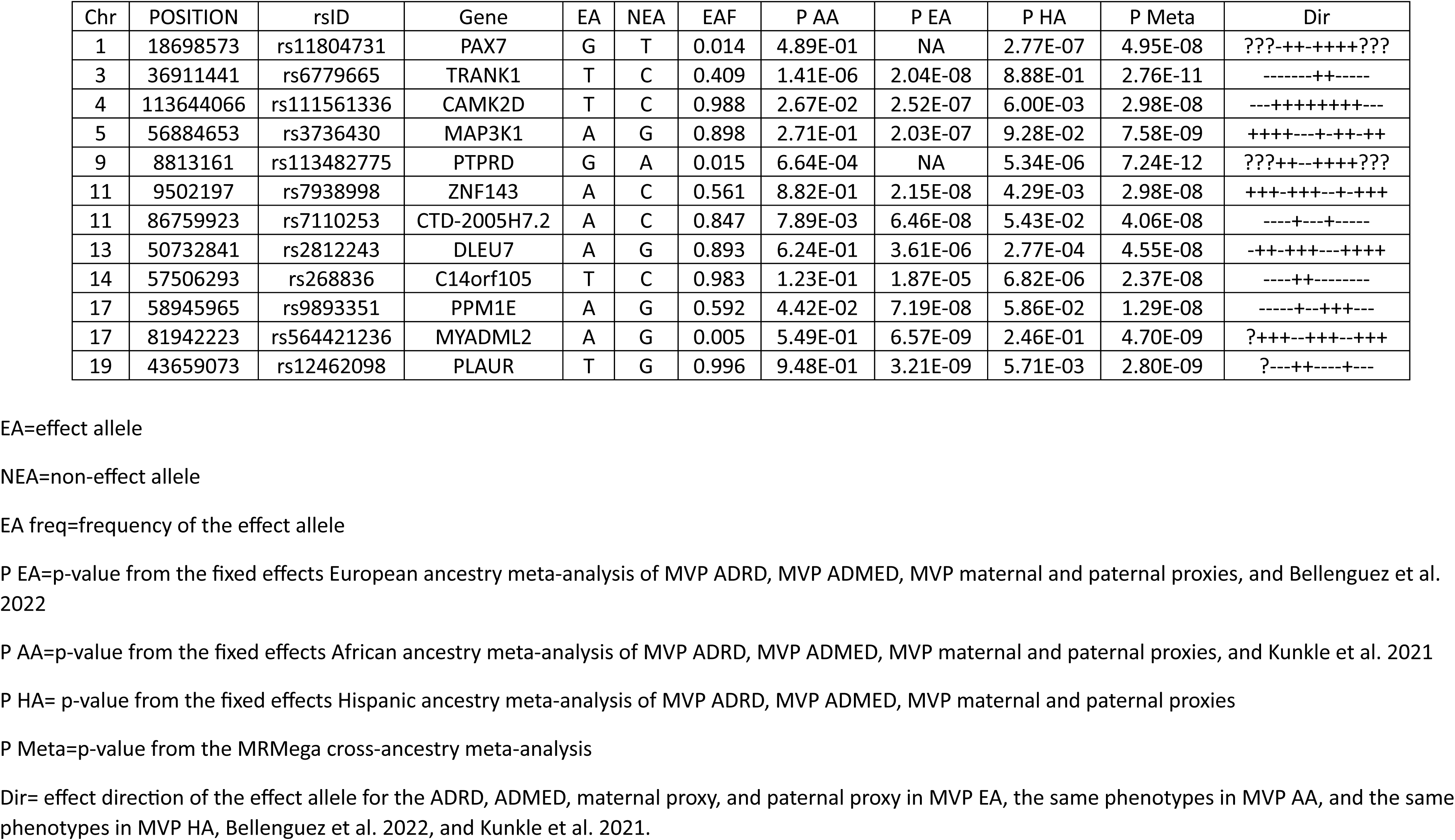
Novel genome-wide significant risk loci in the cross-ancestry meta-analysis of MVP, Bellenguez et al. 2022, and Kunkle et al. 2021.

### Pathway and gene set enrichment analysis

Excluding genes from the *APOE* region, 142 gene sets from 19 categories were significantly enriched across all the meta-analyses conducted. The enrichment included several previously identified pathways/GO terms related to known AD pathology, such as immune regulation, protein degradation, lipid binding, and endosome/lysosome function. Significant enrichment was also observed in microglia-specific genes.. In addition, both the EA and CA meta-analyses showed significant enrichment for association with a largely non-overlapping set of genes in the Parkinson’s disease GWAS catalogue including the novel GWS genes *CAMK2D* and *RERE*. Enrichment was also observed in gene sets from previous GWAS catalogues for cognitive function, schizophrenia, stroke risk, glycemic response to metformin, fruit consumption, personality traits, and several body morphology traits. Several of the other newly GWS genes appear in significantly enriched functional gene sets relevant to ADRD and associated cognitive traits, including several gene sets that contain previously identified AD risk genes. Table S6 shows these results for the MVP EA ancestry meta-analysis, the full EA meta-analysis, and the cross-ancestry meta-analysis, respectively. *FANCA*, *SKA2*, *RERE*, and *PRSS23* appear in gene sets related to eye phenotypes and brain morphology along with *PILRA* and *ZCWPW1*. *LLGL1*, *CAMK2D*, and *TMEM163* appear in gene sets related to vesicular transport. *CR1*, *SORL1*, *MS4A3*, *CR1*, *BIN1*, and *ADAM10*. *PPME1* and *SKA2* are in the GWAS catalogue-defined gene set Cognitive Function.

GARFIELD enrichment analysis indicated that AD-associated SNPs were enriched in several functional and regulatory categories and cell types. Significant enrichment of AD-associated variants within enhancer elements active in K562 erythroleukemic cells (OR = 15.27, P = 3.78x10^-16^) was identified, strongly implicating gene-regulatory functions in white blood cells possibly involving immunoregulation and inflammation as central to the AD’s genetic etiology. Additionally, we identified significant enrichment of AD-associated variants with DNase hypersensitivity in glioblastoma brain cells (OR = 4.89, P = 6.70x10^-7^). Finally, the strongest and most significant enrichment of AD-associated SNPs was in CD19+ human B lymphocytes involved in adaptive immune response, further highlighting an important role of immunoregulation in the genetic architecture of AD (Figure S37).

### Differential expression, eQTL, co-localization, and functional enrichment analysis

In AAs, *ZHX3* was significantly differentially expressed in PFC (OR=3.69, FDR=0.01). In the EA donors, 14 genes were differentially expressed across one or more of the three brain regions (*ZHX3*, *RERE*, *ZNF143*, *CHN2*, and *FERMT2* in AC; *ZHX3*, *ZNF143*, *RERE*, *RASGRP3*, *TRANK1*, *TNIP1*, *SKA2*, *TMEM163*, *PAX7*, *ICA1L*, *PRSS23*, *CHN2*, and *FANCA* in DLPFC; and *CHN2* and *RERE* in PCC) (Table S7). Increased expression of one of these genes (*PAX7*,) was associated with lower ARDR risk with ORs between 0.40 and 0.72. The opposite pattern was observed in the remaining ten genes, with ORs between 1.32 and 3.69.

For the genes within 1 MB of the novel GWS SNP, shown in Tables 2-5, 132 genes representing 156 SNP x expression association pairs showed significant (FDR≤0.05) evidence of eQTL effects of the peak SNP in the EA population (Table S6). The significant eQTL SNPs were associated with both reduced expression in cases (N=91) and increased expression (N=65). There was evidence that four of the GWS SNPs in novel genes (rs117081003 in *RB1*, rs35564151 in *ZNF143*, rs111561336 in *CAMK2D*, and rs35564151 in *TMEM163*) were eQTLs for their respective genes. No significant eQTL effects were identified in the AA brain data.

The co-localization analysis provided additional insight into the SNPs with regulatory effects on nearby genes in several tissues, including SNPs in previously identified genes (*CR1, CLU, TREM2, CD2AP, PILRA, SHARPIN, ACE, CD33, EHHA1, CASS4*). Several of the peak SNPs significantly (posterior probability 4 ≥ 0.8) regulate expression of nearby genes (*AGFG2, AZGP1, LAMTOR4, NYAP1, DCAF7, CSTF1*). Of the novel/newly GWS SNPs, there evidence for co-localization with the SNP in *TRANK1* regulate expression of both *TRANK1* and *LRRFIP2.* Figure S38 shows all SNP x gene colocalization results with posterior probability 4 values ≥ 0.7.

## Discussion

The addition of data from MVP participants to a cross-ancestry ADRD GWAS including previously generated summary statistics from EA and AA cohorts in the U.S. and Europe (Bellenguez et al. 2022. and Kunkle et al. 2021.) constituted one of the largest samples for this purpose and yielded approximately 27 newly GWS risk loci. Notably, this study included the largest number of non-EA individuals, which likely further boosted power for a cross-ancestry GWAS. Although most of the novel loci identified in the cross-ancestry analysis were supported primarily by the EA group, other ancestry groups contributed to the GWS associations for the majority (i.e., *PAX7, CAMK2D, MAP3K1, PTPRD, CTD-2005H7.2, DLEU7, C14orf105, and PPME1*). In fact, most of the evidence for *PAX7* and *PTPRD* was contributed by the AA and HA cohorts. They may also be sex-specific effects, given the vast majority of MVP participants are male. Alternatively, they may have been detected due to the inclusion of non-AD dementia cases, since only the variants in *RB1*, *FERMT2*, *SKA2*, and *PPM1E* showed nominally significant associations with diagnosed AD in the ADGC EA cohorts^31^.

Our study supports the relevance of pathways identified in prior studies, including immune regulation, microglia, and peripheral immune cells^32-34^. We also found enrichment for AD-associated genes involved in lipid binding^35^, despite the exclusion of the *APOE* region from these analyses. In addition, findings from analyses of the EA and combined ancestry groups showed significant enrichment for a largely non-overlapping set of genes associated with Parkinson’s disease (PD).

Several genes in the novel GWS loci have been implicated in AD or ADRD-relevant biology. *CHN2* is a Rac1-specific guanosine triphosphate GTPase-activating protein involved in neuronal development and axon pruning that has been previously associated with AD in a GWAS of individuals with high genetic risk for AD in the UKBB^36^ and with risk of long COVID-19^37^. *IPO7* encodes a nuclear pore import receptor protein that transports three ribosomal subunits across the nuclear membrane^38^. GAL is brain neuropeptide regulating synaptic transmission, glutamate release, and potassium channels that has been reported to have a neuroprotective action by attenuating b-amyloid-induced loss of neurons. GAL has an anti-neuroinflammation action, and its overexpression was associated with inhibited cognitive functions in a mouse model^39^. SEC13 regulates numerous immune functions through a role in golgi function^40^. *TMEM163* is a zinc transporter protein involved in synaptic vesicle transport, zinc homeostasis in oligodendrocyte development, myelin formation^41^, and endosomal function^42^. *TMEM163* variants have been associated with early-onset PD^43^. Proteome-wide association studies found associations of ICAL1 abundance in the brain with AD^44^, as well as small vessel strokes and intracerebral hemorrhages^45^. TRANK1 has roles in pathways regulating synaptic function, dendritic spine density, circadian rhythms, and calcium signaling, and may be a risk gene for bipolar disorder^46^. *TNIP1* is involved in immune system regulation through nuclear factor kappa-B and has been implicated in several autoimmune disorders^47^ and possibly ALS^48^. One study reported that expression of *PRSS23,* a serine protease in the trypsin family, differed significantly between AD cases and controls, and may influence AD risk through a mechanism involving CD8+ T-cells expressing lower levels of IL-7 receptor alpha^49^. *ZNF143* regulates transcription of *AP2S1*, a protein that regulates APP degradation through late endosome-lysosome fusion in mice^50^. *SKA2* has been implicated in AD pathology through neuroinflammation and secretory autophagy^51^ and possibly stress signaling mediated by the glucocorticoid receptor pathway and hypothalamic–pituitary–adrenal axis^52^. *ZHX3* is a transcriptional repressor and was one of the top five upregulated genes in a study comparing AD cases and controls^53^. There is an extensive literature linking *CAMK2D* to AD-associated pathology (reviewed in^54^) through synaptic plasticity and learning^55^, tau phosphorylation^56^, and amyloid beta^57^. Similarly, there is a growing body of evidence linking MAPK signaling to AD^58,59^ and a miRNA targeting MAP3K1 has been shown to inactivate MAPK signaling, suggesting it may be a promising target for downregulation as an AD therapy^60^.

The DGE and eQTL results provide further context to the potential functional impact of the novel SNP associations and ADRD pathology in general. The associations in *ZNF143* and *TMEM163* represent novel genes with evidence that differential expression affects ADRD risk and that the specific SNPs identified drive expression. The co-localization results indicate *TRANK1* may also be under regulatory control by the peak SNP. This, combined with their ADRD-relevant functions, make them potential candidates for treatment targets.

## Limitations

Our use of proxy cases, AD medication cases, and ADRD instead of a strict validated AD diagnosis leaves open the possibility that our newly associated loci are general dementia risk factors rather than AD specifically. Further study will be needed to determine whether these are AD specific, more general dementia loci, or risk loci for a related dementia. Additionally, our eQTL analyses were limited in terms of the tissues examined, and causal effects may be observable in other brain regions. There was also moderate inflation in the EA and CA meta-analysis summary statistics. This is not uncommon in large GWAS meta-analyses and likely reflects polygenicity rather than systematic issues with the models. Finally, we acknowledge that the inclusion of Asian ancestry cohorts would have improved the global representativeness of this project, but we were not able to include an Asian ancestry meta-analysis in MVP because of the small number of Asian-ancestry MVP participants.

## Conclusions

In this study, we have presented analyses from a large biobank comprised of US Veteran volunteers. While the accuracy of AD/ADRD diagnoses based on electronic medical record (EMR) data is less than clinical examination by experts, MVP enables opportunities for testing hypotheses that are not possible in cross-sectionally ascertained cohorts. Specifically, retrospective follow back health history for most MVP participants allows for the examination of the contribution of comorbidities, health factors, and environmental exposures to AD/ADRD pathogenesis, and how these factors interact with AD genetic risk factors (e.g. ^61,62^). Further coordinated analyses involving data from MVP and international AD consortia are likely to yield greater insight into the genetics of ADRD and the degree to which ADRD risk factors generalize across populations.

## Methods

### MVP Cohort description

MVP is a nationwide research program established in 2011 as part of a health initiative to identify Veteran-relevant disease risk factors^63,64^. Upon enrolment, MVP participants provide a blood sample and consent to the access of their VA electronic medical record for research purposes. The VA EMR is the primary source of phenotype data, including treatment codes and prescription history, obtained from MVP participants over a period of as long as 30 years. Many MVP participants completed surveys that collect many types of data, including nutrition, health factors, family history, military service, and psychiatric-disorder history^65^. The fourth MVP genotype release includes ∼650K MVP participants (methods described below).

### Dementia Diagnosis and Classification

While our primary GWAS is a case/control analysis of ADRD in MVP data, we also conducted paternal and maternal proxy GWASs, which have been shown to yield reliable results for ADRD. In addition, we performed an AD “medication” GWAS, because we previously demonstrated that adding cases identified based on usage medications for AD to ICD-code identified cases resulted in stronger associations with APOE genotypes and an AD PRS^66^. However, in light of the limitations of the EMR in terms of specificity of treatment coding, we focused on a broader case definition of ADRD.

1. **<u>ADRD</u>:** ADRD cases were defined as participants with diagnosis codes for AD, non-specific dementia^67^, or other diagnosis codes for dementias that co-occur and/or have symptom overlap with AD (i.e., vascular dementia, Lewy body dementia, frontotemporal dementia^68^). Two or more of these diagnosis codes and an age of onset (first diagnosis code) at or above age 60 were requirements for inclusion as an ADRD case. Because AD accounts for more than 50% of all dementia diagnoses^69^ and due to VA code usage^67^, it is likely that a large portion of MVP ADRD cases have AD. This conclusion is supported by our recent study^66^ showing that APOE ɛ4 genotype and an AD genetic risk score were more strongly associated with the ADRD diagnosis assigned based on the algorithm for ADRD using ICD codes than a stricter AD algorithm that only included AD diagnosis codes.
2. **<u>Dementia Medication Prescription</u>:** MVP participants who did not meet ADRD criteria described above were classified as an ADRD “medication” case if the VA EMR included a prescription of any AD medication, specifically cholinesterase inhibitors (e.g., donepezil, galantamine, rivastigmine or memantine. ADRD defined in this manner is also associated with *APOE* ɛ4 genotype and an AD genetic risk score^66^.
3. **<u>Parental Proxy Classification</u>:** Most MVP participants completed the “baseline” survey that included questions regarding a wide variety of topics including employment, personality, psychiatric disorders, and family history of disease. Excluding individuals classified as ADRD using any of the above criteria, participants who reported a history of “Alzheimer’s/ Other dementia” in their father or mother were classified as paternal or maternal proxy AD cases, respectively, and individuals with two affected parents were classified as paternal proxy cases to maintain independence of paternal and maternal proxy AD GWASs and minimize sample size disparities.
4. **<u>Control Definitions</u>:** Independent groups of controls were used for the ADRD and medication ADRD GWASs. Controls were 65 years of age or older at their last visit in a VA clinic, had no ICD codes for any dementia-related disorder (not limited to the ICD codes defining ADRD) or MCI (see ^66^ for details), did not report having a parent with dementia, and had no history of AD-medication prescription. The control sets for the ancestry-specific ADRD analyses were selected first, randomly, in numbers equal to four times the number of ADRD cases. Four times the number of AD medication cases within each ancestry group were then randomly selected from the remaining control pool. The remaining controls were randomly split between the paternal and maternal proxy analyses. In addition, proxy GWAS controls were also added from MVP participants who were at least 40-65 years old at baseline and reported no parental dementia, because their parents would presumably have had a relatively low risk for dementia.

### Genotype Data Generation, Processing and Imputation, and Relationship Checks and Principal Components Analysis

Genotype data processing and cleaning was performed by the MVP Bioinformatics core. The genotype data were generated using the MVP 1.0 custom Axiom array which assays 668,418 markers^63^. Quality control (QC) included checks for sex concordance, advanced genotyping batch correction, and assessment for relatedness^63^. The chip design and genotype cleaning pipeline have been described elsewhere^63^. The MVP Phase 4 genotype data release includes imputed genotype data for 62 million variants assessed for approximately 650,000 subjects. Imputation was performed using the TOPMed^70^ imputation panel. Prior to imputation, SNPs with high missingness (20%), monomorphic, not in Hardy-Weinberg equilibrium, or that differed in frequency between batches were removed. Phasing was done with SHAPEIT4 v 4.1.3 and imputation was performed with MINIMAC4. One of each pair of related individuals, defined as a kinship coefficient of 0.09375 or higher, were removed from analysis using a scheme that prioritized a case over a control while the older control was selected if both were controls with no consideration for whether they had an AD medication or parental proxy diagnosis. When both members of a pair were ADRD cases, we selected a subject with AD-specific ICD codes or, in the absence of ICD codes, one individual was randomly selected.

Principal components of ancestry (PCs) were computed with FlashPCA2^71^ within the HARE^30^-defined AA, EA, and HA groups using linkage-disequilibrium pruned sets of SNPs that excluded the major histocompatibility complex region of chromosome 6. *APOE* genotypes were determined using the “best guess” imputed genotypes (80% confidence threshold) for the rs7412 and rs429358 SNPs which were well imputed in the three ancestry groups (r^2^ > 85%).

### GWAS analysis methods

A separate GWAS was performed for ADRD determined by ICD code and medication use, and maternal and paternal proxy AD. Analyses were performed separately in each ancestry group. The association of AD outcomes with each SNP having a minor allele frequency ≥ 0.001% and imputation r^2^ ≥ 0.4with AD outcomes was evaluated using logistic regression models implemented in Plink 2^72^ including covariates for sex and the first 10 ancestry-specific PCs. Age was not included since cases and controls were selected based on age. Since the effects of genes in the *APOE* region have been studied extensively and these genes would still appear in any pathways that contain them, SNPs in that region on chromosome 19 (HG38 genomic position 43,904,887-45,909,392 bp) were excluded from all downstream analyses.

Results within ancestry groups were combined by meta-analysis using a fixed effects, inverse variance weighted model implemented in ^73^. A cross-ancestry (CA) meta-analysis was conducted using Meta-Regression of Multi-AncEstry Genetic Association (MR-MEGA)^74^. Summary results obtained from large AD GWAS conducted in independent data sets of EA^4^ and AA^75^ individuals were combined in the respective MVP ancestry groups by meta-analysis. The effect sizes and standard errors for SNPs in the proxy AD GWAS summary statistics were both multiplied by two to correct for attenuation bias caused by measurement error^10^.

### Annotation

Gene mapping, functional annotation, and pathway/gene set enrichment analysis were performed using the FUMA GWAS software^76^. We designated a GWS associations as novel if it was ≥ 1 Mb from a peak SNP in a previously reported locus ^4,29,77^, or between 250 Kb and 1 Mb away from, but with low LD with (pairwise r^2^ ≤ 0.2 in EA), a previously identified peak SNP. The LD coefficient was calculated using the LDlinkR package (https://cran.r-project.org/web/packages/LDlinkR/vignettes/LDlinkR.html) with the Phase 3 (Version 5) 1,000 Genomes Project reference table and, where possible, a within ancestry reference panel. In the CA analysis, we utilized the EA reference panel to provide a conservative approach to identification of novel variants. This resulted in cases where we report a novel GWAS SNP that is near, but in low LD with a previously reported genetic locus. We retained these to emphasize that some of these loci may harbor multiple causal risk variants.

### Differential expression, eQTL, co-localization, and functional enrichment analysis

RNA-seq data derived from brain tissue obtained from 433 EA participants of the Religious Orders Study and Memory and Aging Project (ROSMAP) cohort and 177 AA participants s from 14 Alzheimer’s Disease Research Centers (ADRCs) across the United States^78^ were used for differential gene expression (DGE) analysis of the novel GWS genes and for expression quantitative trait locus (eQTL) analysis of the novel SNPs. Data from both cohorts were processed using the same analysis pipeline. Raw RNA-seq data from anterior cingulate cortex (AC; n=283), posterior cingulate cortex (PCC; n=217) and dorsolateral prefrontal cortex (DLPFC; n=419) in ROSMAP and from prefrontal cortex (PFC) in the ADRC participants were first scaled by transcripts-per-million (TPM) and then adjusted across libraries using the trimmed mean of M-values (TMM) method^79^. DGE analysis was done using logistic regression models with AD as the outcome and normalized expression levels of the novel genes, adjusted for age, sex, and cell type proportions as predictors.

eQTL analysis was done using the Matrix eQTL package in R^80^ to test whether novel SNPs associated with ADRD risk were also associated with expression levels of nearby genes. Genotype data for the ROSMAP individuals were obtained from the Alzheimer’s Disease Sequencing Project (ADSP) Release 4 (R4) whole genome sequencing dataset^81^. Genotypes for the AA cohort were imputed with the 1000 Genomes Phase 3 reference panel filtered for SNPs with hard-calls at ≥80% certainty and R² > 0.2. For each SNP, we tested cis-eQTL effects on genes within 1 MB by adjusting for age, sex, and estimated proportions of six cell types: glutamatergic (GLU), GABAergic (GAB), astrocytes (AST), microglia (MIC), oligodendrocytes (ODC), and endothelial cells (END). Cell type proportions were estimated using a deconvolution method based on expression profiles from single cell RNA-seq data in brain according to the method described by O’Neill et al^82^. Results for both DGE and eQTL analyses were corrected for multiple testing using the Benjamini-Hochberg false discovery rate (FDR) method in R.

Colocalization analysis was performed using coloc.abf from the R package coloc^83^, which applies an approximate Bayes factor framework to estimate the posterior probability that two association signals in a locus share a single causal variant. We tested colocalization between summary statistics from the EA and CA ADRD meta-analyses and cis-eQTL summary statistics from GTEx^84^, as well as plasma pQTL summary statistics from the deCODE SomaScan and UK Biobank Olink studies^85^. These analyses were used to prioritize genes whose genetically regulated expression/protein abundance may underlie ADRD association signals We performed a functional enrichment analysis of our EA fixed effects meta-analysis GWAS summary statistics using GARFIELD^86^. GARFIELD leverages GWAS summary statistics and regulatory/functional annotations in various cells and tissues to identify the characteristics relevant to a trait of interest under different GWAS P-value thresholds^87^. Due to the large number of SNPs obtaining small P values, we used the following thresholds: 10^-5^, 5x10^-8^, 5x10^-10^, and 5x10^-12^. We assessed the significance of SNP enrichment using a P value threshold of 0.0001. Enrichment effect sizes are reported using odds ratios (OR).

## Data Availability

Summary statistics for MVP subgroup and meta-analyses will be deposited in dbGaP under study accession phs001672. Bellenguez et al. GWAS meta-analysis summary results are available at the European Bioinformatics Institute GWAS Catalog (https://www.ebi.ac.uk/gwas/) under accession no. GCST90027158. The Kunkle et al. AA AD GWAS summary statistics are available at NIAGADS DSS under accession number NG00100.

## Acknowledgements

**MVP Cognitive Decline and Dementia during Aging Working Group Members:** Mark W. Logue Chair, Richard L. Hauger Co-Chair, Matthew Panizzon Secretary, Victoria Merrit Founding Member. Members: Julie Lynch, Rui Zhang, Richard Sherva, Lindsay Farrer, William Kremen, Kelly Cho, Jesse Mez, Erika Wolf, Kelly Harrington, Catherine Chanfreau, Yuk-Lam Ho, Debby Tsuang, Qing Zeng, David Salat, David Marra, Mark Miller, Zoe Neale, Jennifer Fonda, Lauren Loeffel, Caroline Nievergelt, Royce Clifford, Sophia Lee, Margaret Gillis, Meghan Wilkinson, Francesca Lopz, Adam Maihofer, Karl Brown.

**VA Million Veteran Program**

**Core Acknowledgements for Publications**

**October 2025**

**MVP Program Office**

Sumitra Muralidhar, Ph.D., Program Director

US Department of Veterans Affairs, 810 Vermont Avenue NW, Washington, DC 20420

Jennifer Moser, Ph.D., Associate Director, Scientific Programs

US Department of Veterans Affairs, 810 Vermont Avenue NW, Washington, DC 20420

Jennifer E. Deen, B.S., Associate Director, Cohort & Public Relations

US Department of Veterans Affairs, 810 Vermont Avenue NW, Washington, DC 20420

**MVP Steering Committee**

Co-Chair: Philip S. Tsao, Ph.D.

VA Palo Alto Health Care System, 3801 Miranda Avenue, Palo Alto, CA 94304

Co-Chair: Sumitra Muralidhar, Ph.D.

US Department of Veterans Affairs, 810 Vermont Avenue NW, Washington, DC 20420

J. Michael Gaziano, M.D., M.P.H.

VA Boston Healthcare System, 150 S. Huntington Avenue, Boston, MA 02130

Adriana Hung, M.D., M.P.H.,

VA Tennessee Valley Healthcare System, 1310 24th Avenue, South Nashville, TN 37212

Dave Oslin, M.D.

Philadelphia VA Medical Center, 3900 Woodland Avenue, Philadelphia, PA 19104

Deepak Voora, M.D.

Durham VA Medical Center, 508 Fulton Street, Durham, NC 27705

**MVP Co-Principal Investigators**

J. Michael Gaziano, M.D., M.P.H.

VA Boston Healthcare System, 150 S. Huntington Avenue, Boston, MA 02130

Philip S. Tsao, Ph.D.

VA Palo Alto Health Care System, 3801 Miranda Avenue, Palo Alto, CA 94304

**MVP Core Operations**

Jessica V. Brewer, M.P.H., Director, MVP Cohort Operations

VA Boston Healthcare System, 150 S. Huntington Avenue, Boston, MA 02130

Mary T. Brophy M.D., M.P.H., Director, VA Central Biorepository

VA Boston Healthcare System, 150 S. Huntington Avenue, Boston, MA 02130

Kelly Cho, M.P.H, Ph.D., Director, MVP Phenomics

VA Boston Healthcare System, 150 S. Huntington Avenue, Boston, MA 02130

Lori Churby, B.S., Director, MVP Regulatory Affairs

VA Palo Alto Health Care System, 3801 Miranda Avenue, Palo Alto, CA 94304

Jacob T. Kean, Ph.D., Acting Director, VA Informatics and Computing Infrastructure (VINCI)

VA Salt Lake City Health Care System, 500 Foothill Drive, Salt Lake City, UT 84148

Saiju Pyarajan Ph.D., Director, Data and Computational Sciences

VA Salt Lake City Health Care System, 500 Foothill Drive, Salt Lake City, UT

Robert Ringer, Pharm.D., Director, VA Albuquerque Central Biorepository

VA Boston Healthcare System, 150 S. Huntington Avenue, Boston, MA 02130

Luis E. Selva, Ph.D., Director, MVP Biorepository Coordination

New Mexico VA Health Care System, 1501 San Pedro Drive SE, Albuquerque, NM 87108

Shahpoor (Alex) Shayan, M.S., Director, MVP PRE Informatics

VA Boston Healthcare System, 150 S. Huntington Avenue, Boston, MA 02130

Brady Stephens, M.S., Principal Investigator, MVP Information Center

VA Boston Healthcare System, 150 S. Huntington Avenue, Boston, MA 02130

Stacey B. Whitbourne, Ph.D., Director, MVP Cohort Development and Management

Canandaigua VA Medical Center, 400 Fort Hill Avenue, Canandaigua, NY 14424

