## Supplementary Figures for "A Genome-wide Association Study of Alzheimer’s Disease and Dementia in a Large Multi-ancestry Military Cohort Identifies Many New Dementia-Associated Loci"

Figure S1. CHNS in MVP EA

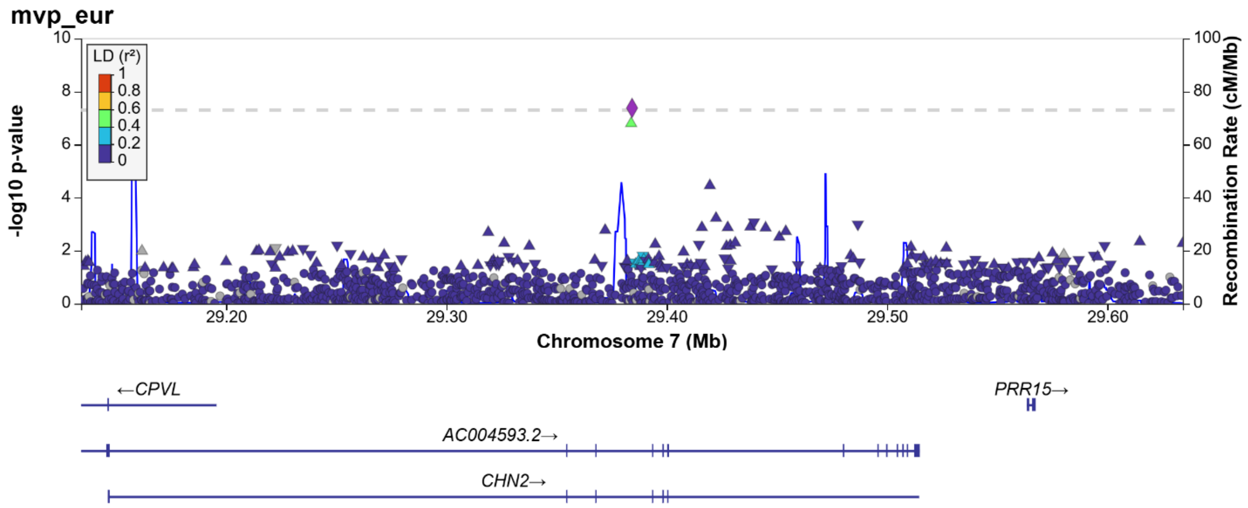


Figure S2. IPO7 in MVP EA

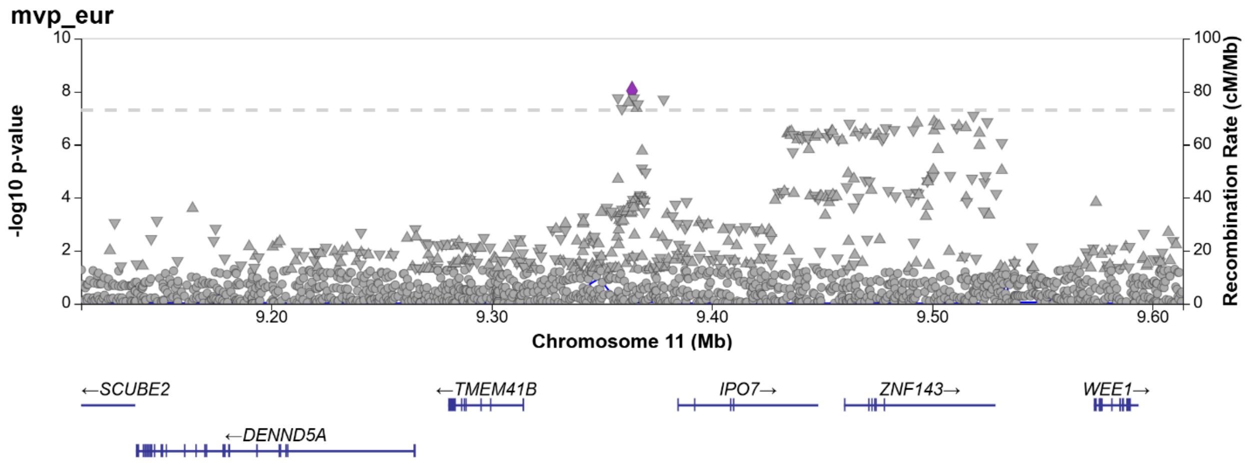

Figure S3. GAL in MVP EA

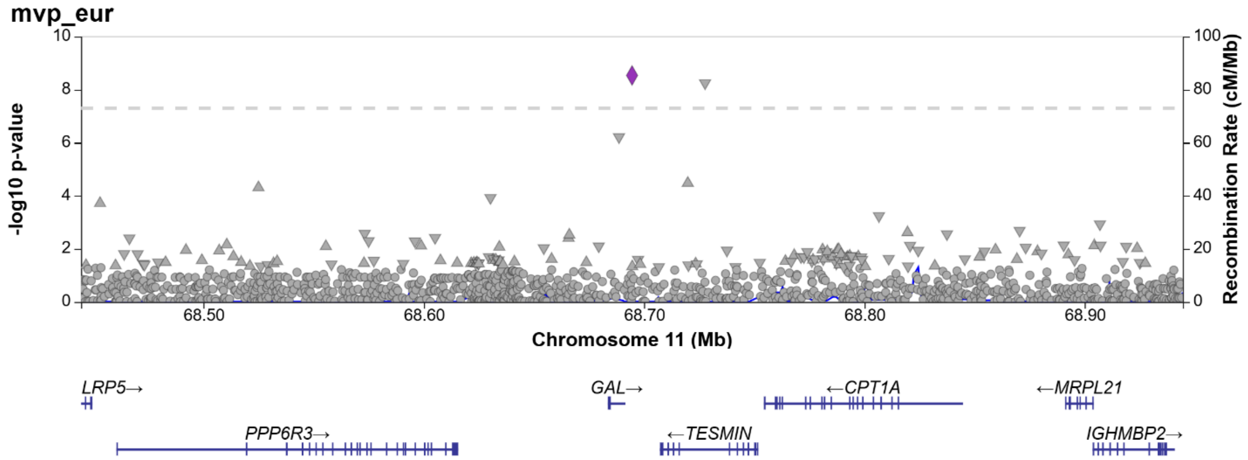


Figure S4. RB1 in MVP EA

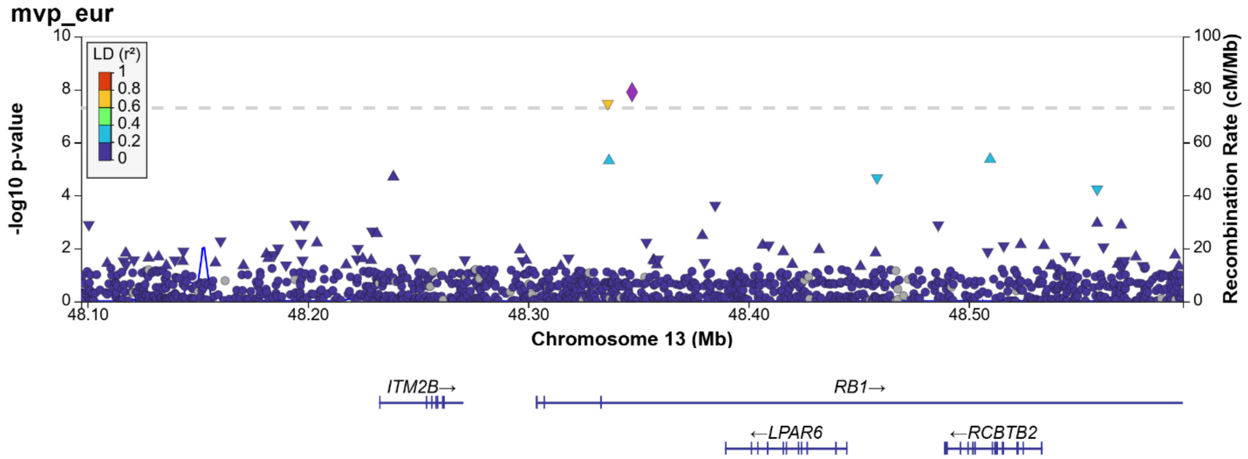

Figure S5. PYCR1/MYADML2 in MVP EA

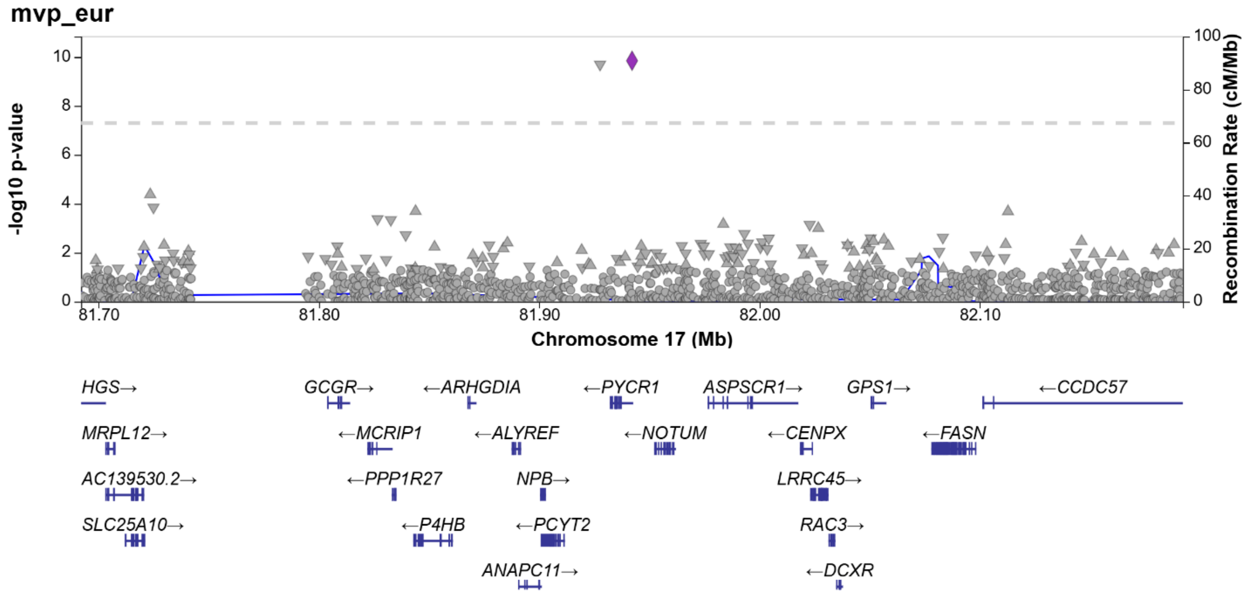


Figure S6. RERE in all EA

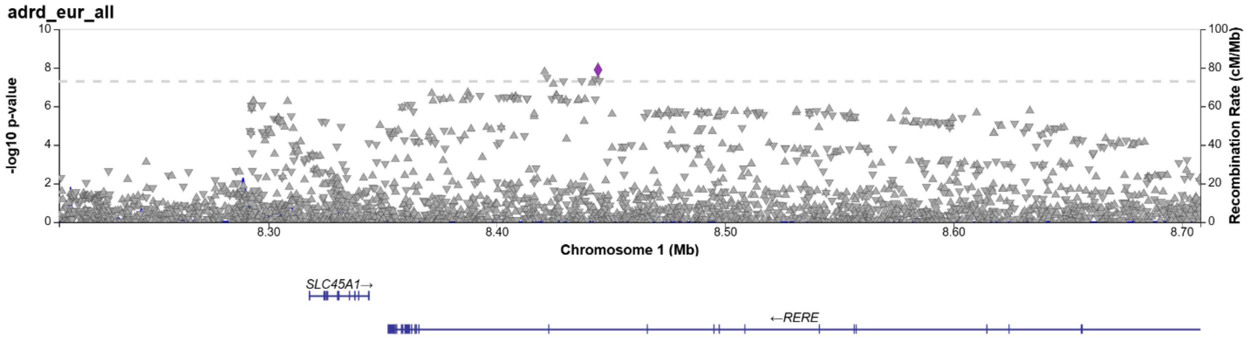

Figure S7. TRANK1 in all EA

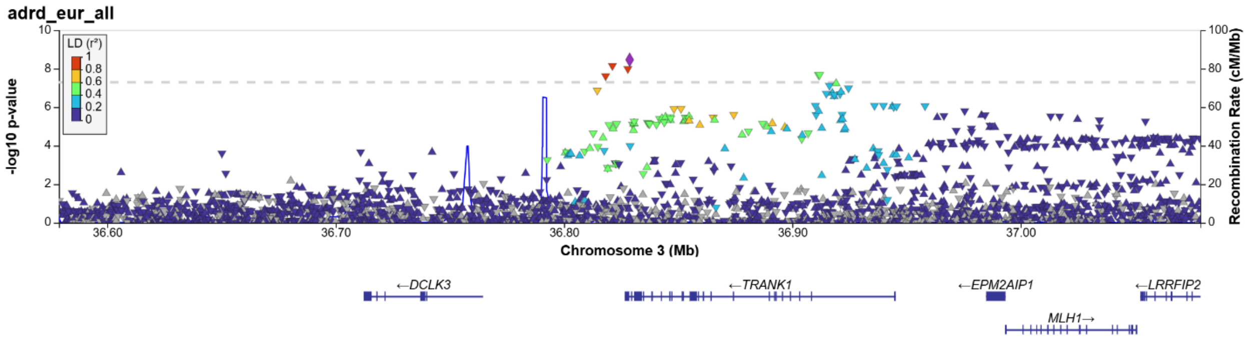


Figure S8. CAST in all EA (SNP in not in gene on Locuszoom, is in NCBI)

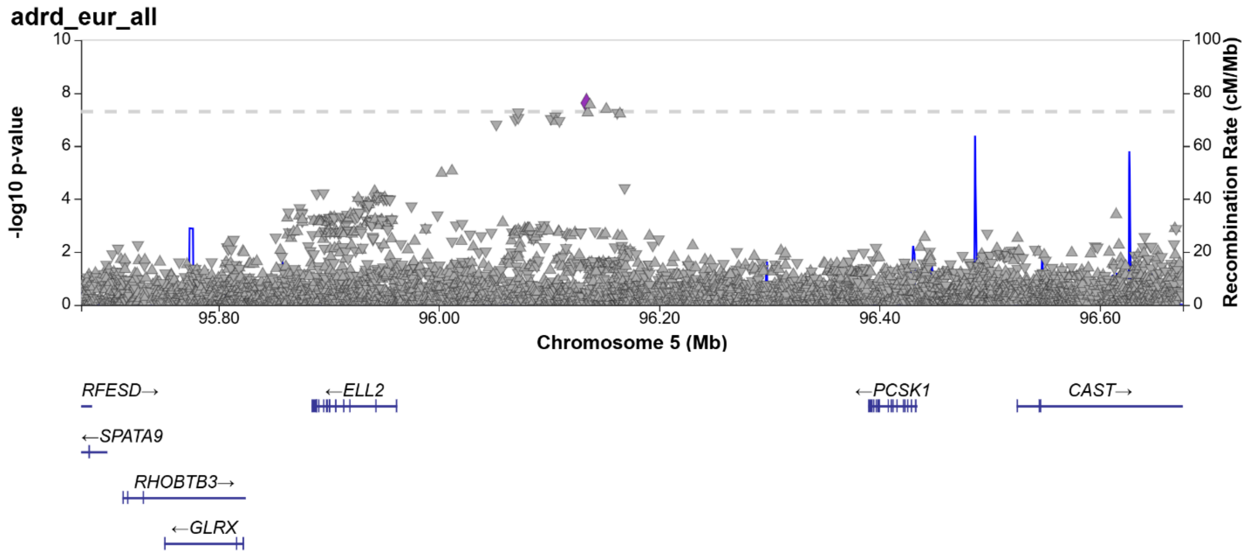

Figure S9. UBTD1in all EA

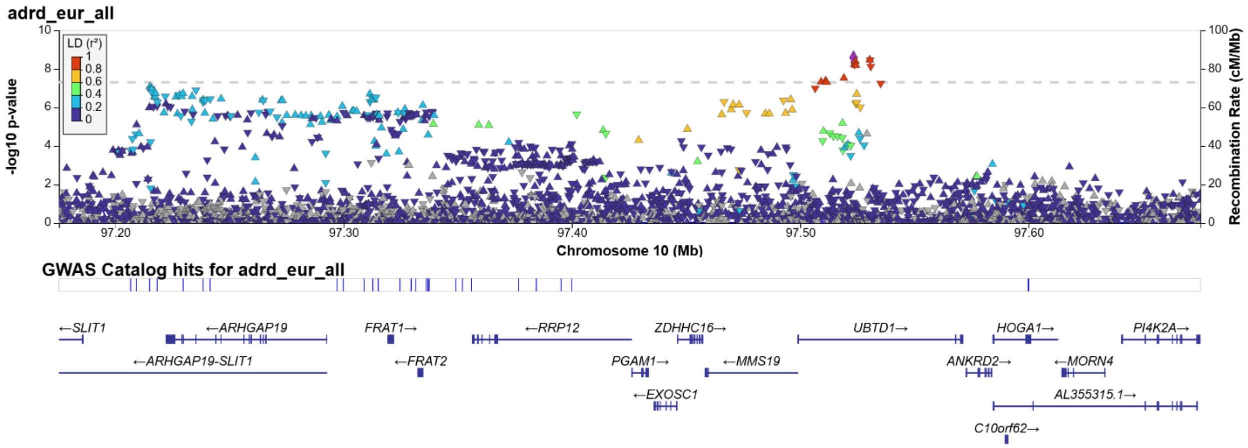


Figure S10. ZNF143 in all EA

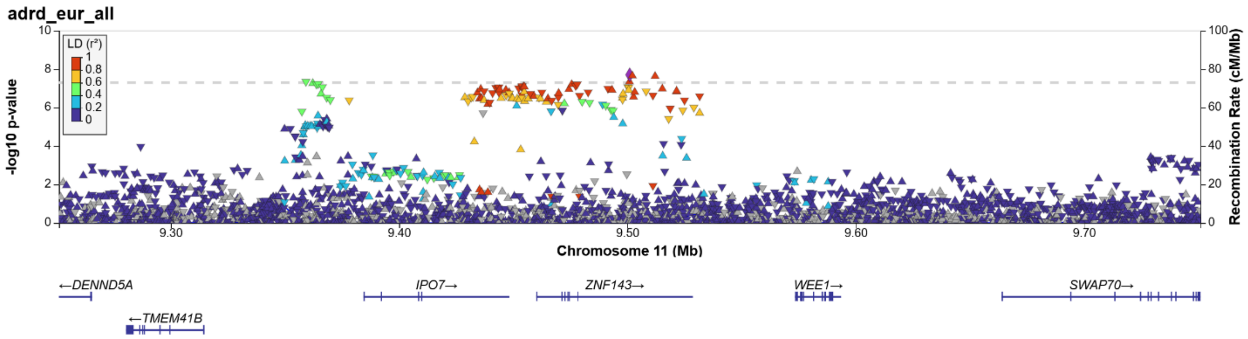

Figure S11. PRSS23 in all EA

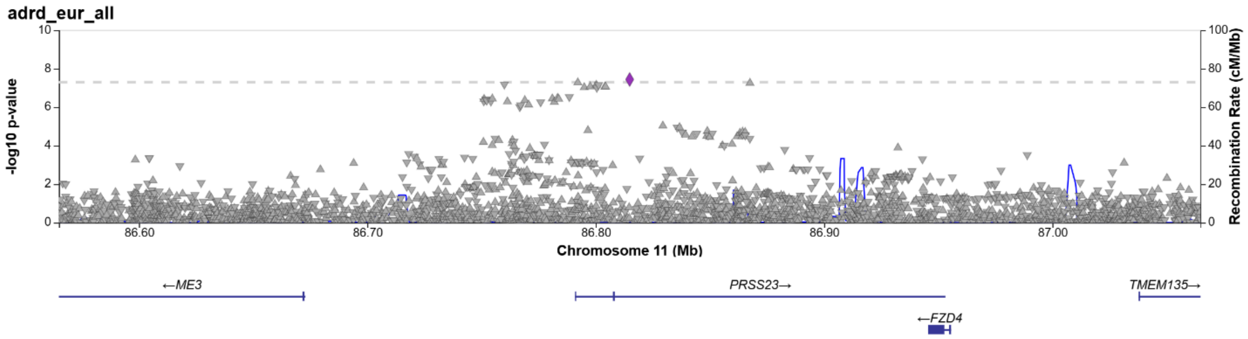


Figure S12. FERMT2 in all EA

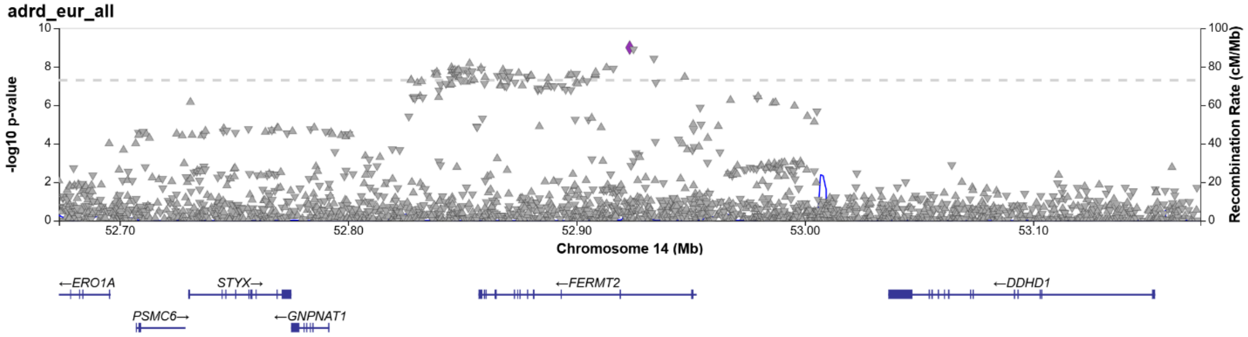

Figure S13. SKA2 in all EA

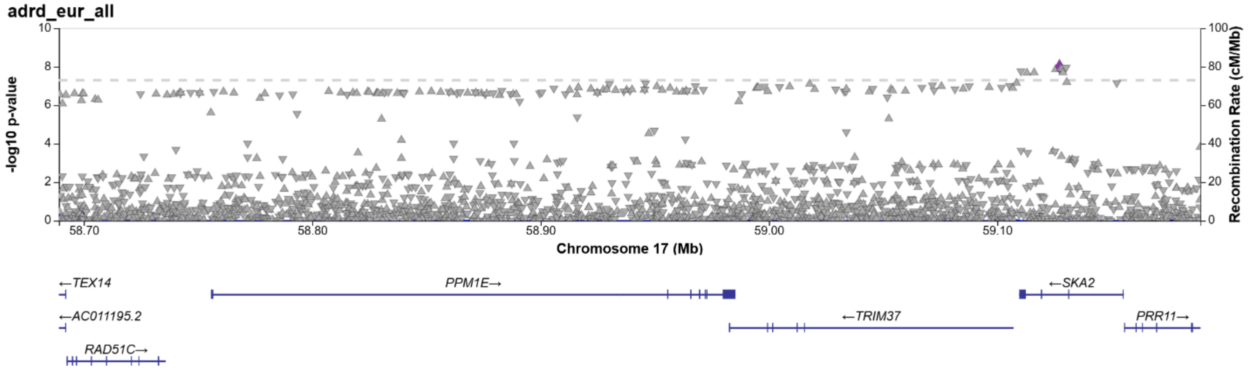


Figure S14. MYADAML2 in all EA

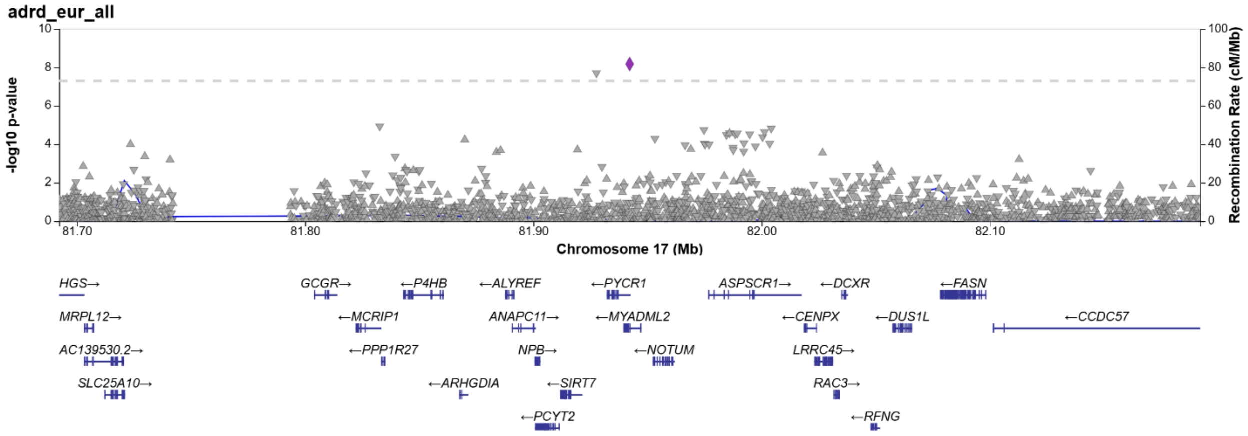

Figure S15. PLAUR in all EA

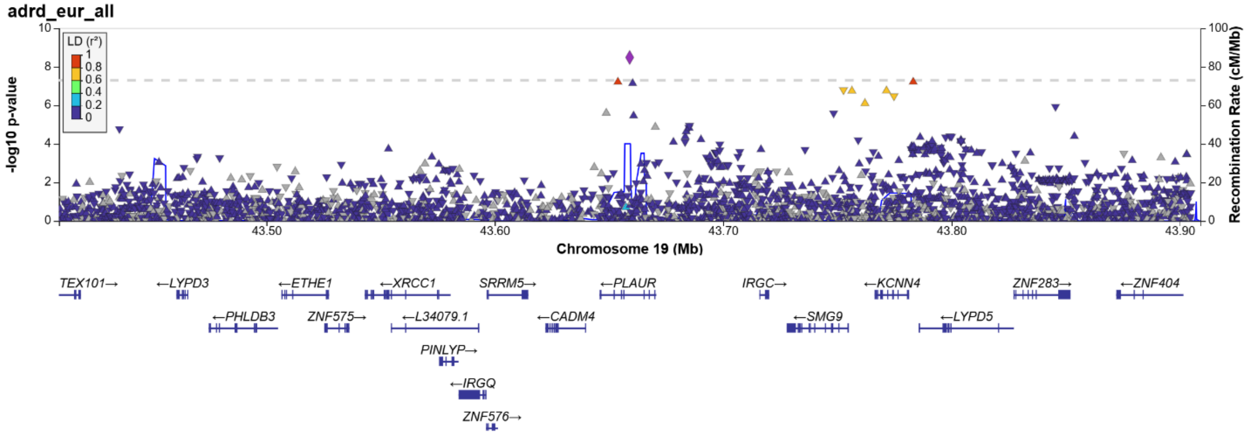


Figure S16. ZHX3 in all EA

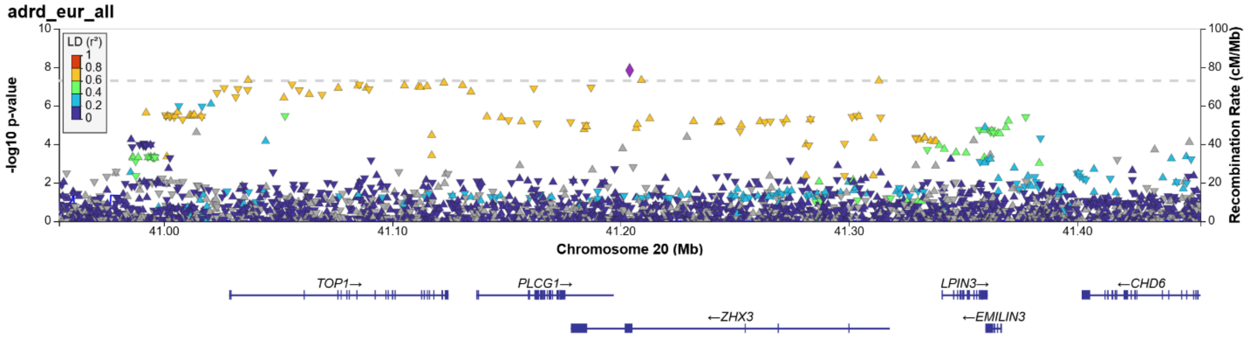

Figure S17. RASGRP3 in all AA

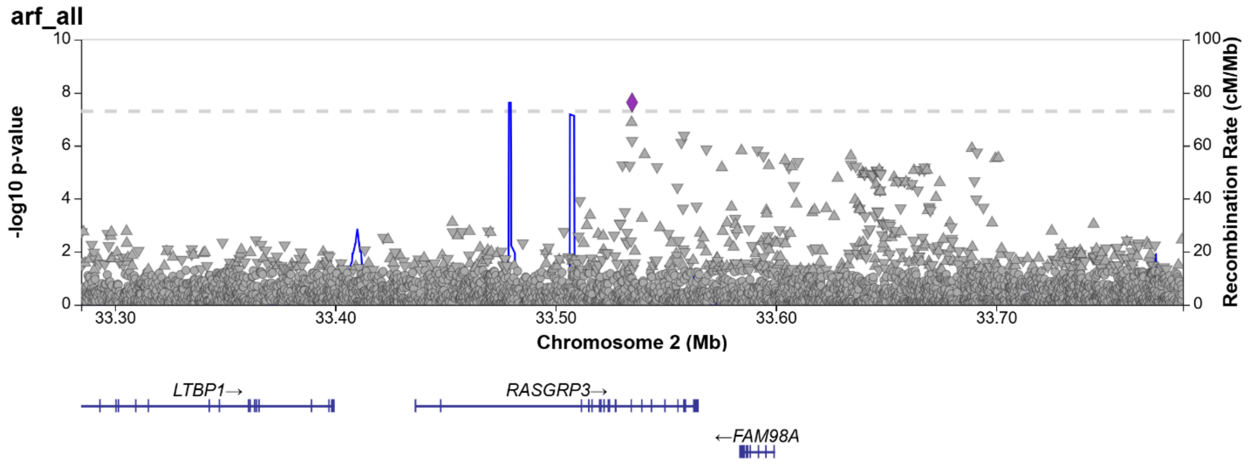


Figure S18. SEC13 in all AA

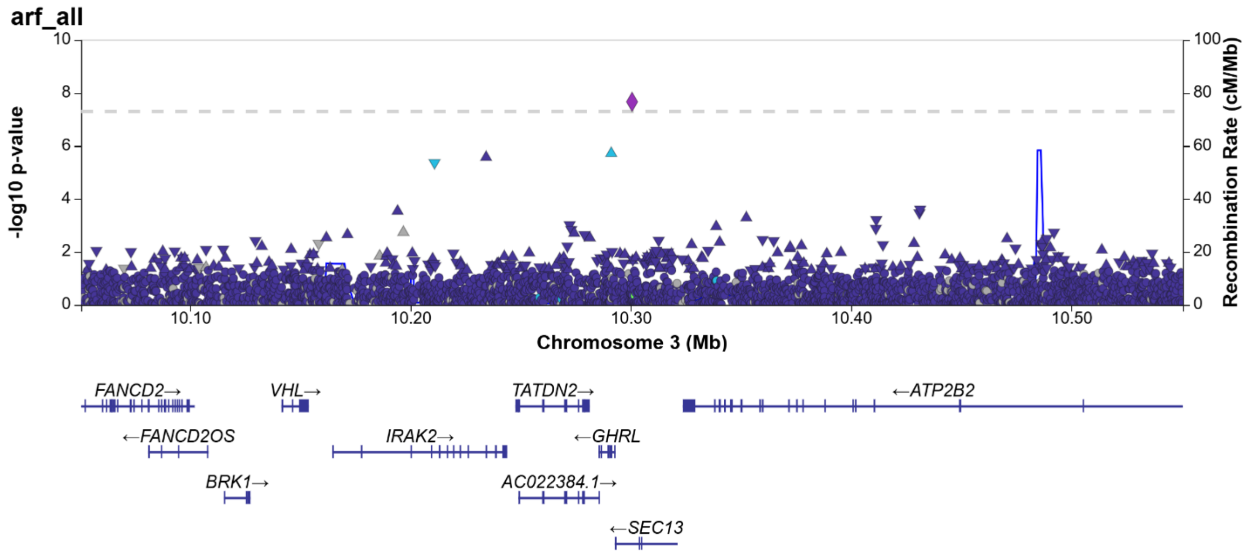

Figure S19. COMMD10 in MVP HA

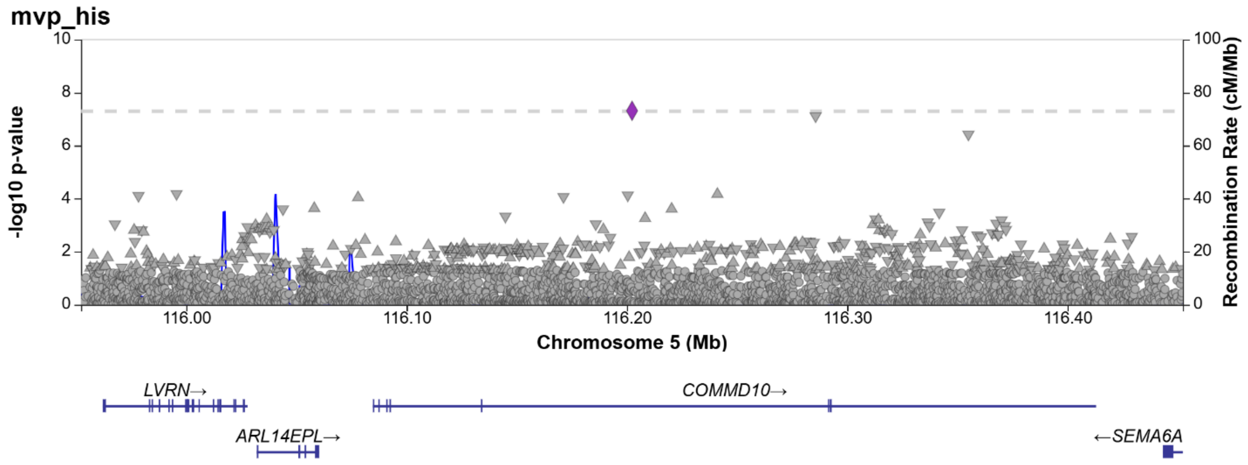


Figure S20. CALCR in MVP HA

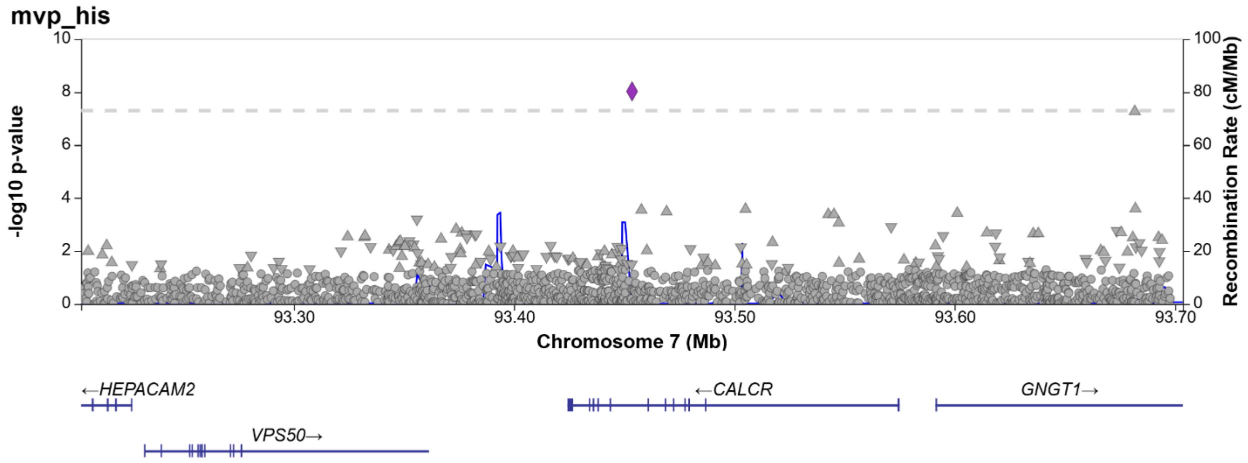

Figure S21. BRINP1 in MVP HA

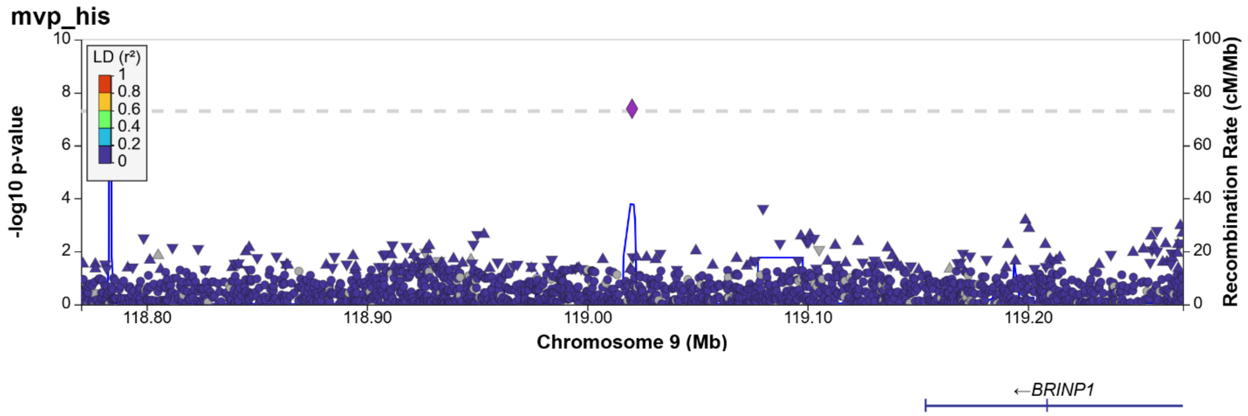


S22. NAP1L1-LOC641695 in MVP HA

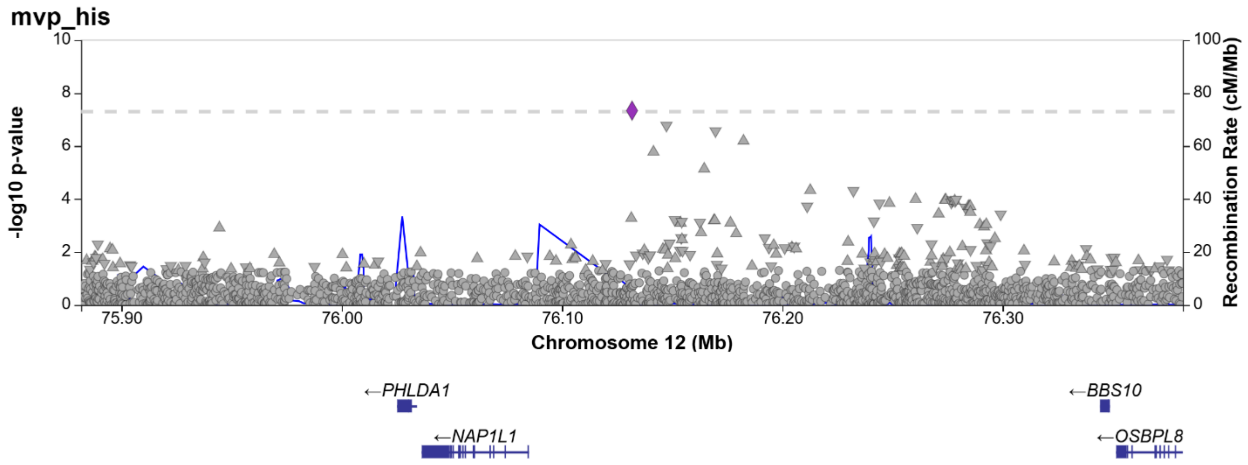

Figure S23. NUFIP2-TAOK1 in MVP HA

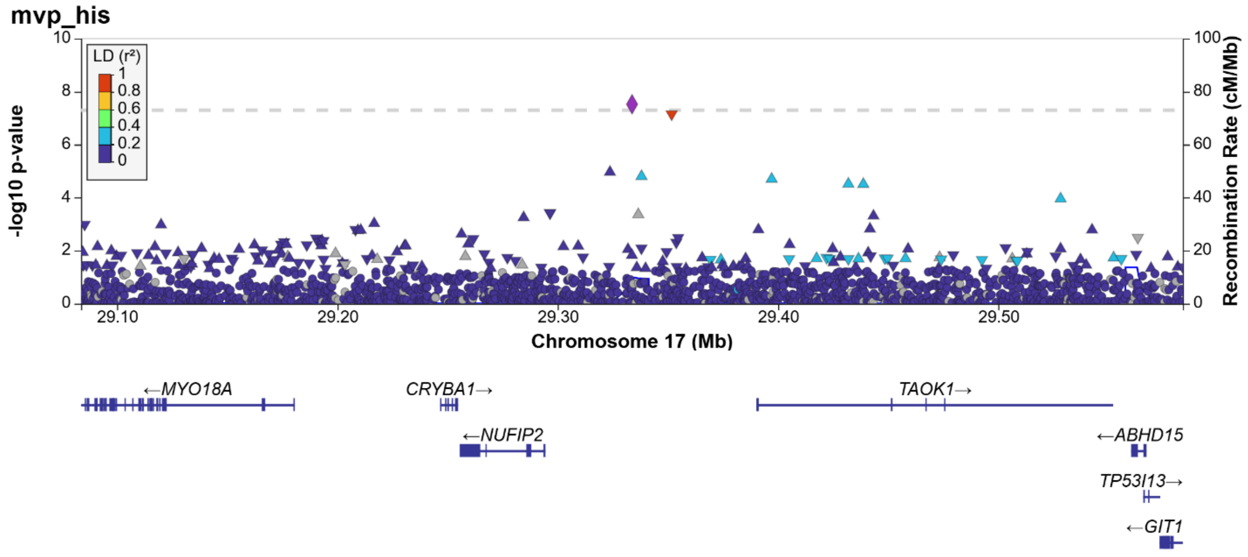


Figure S24. ZSCAN5A in MVP HA

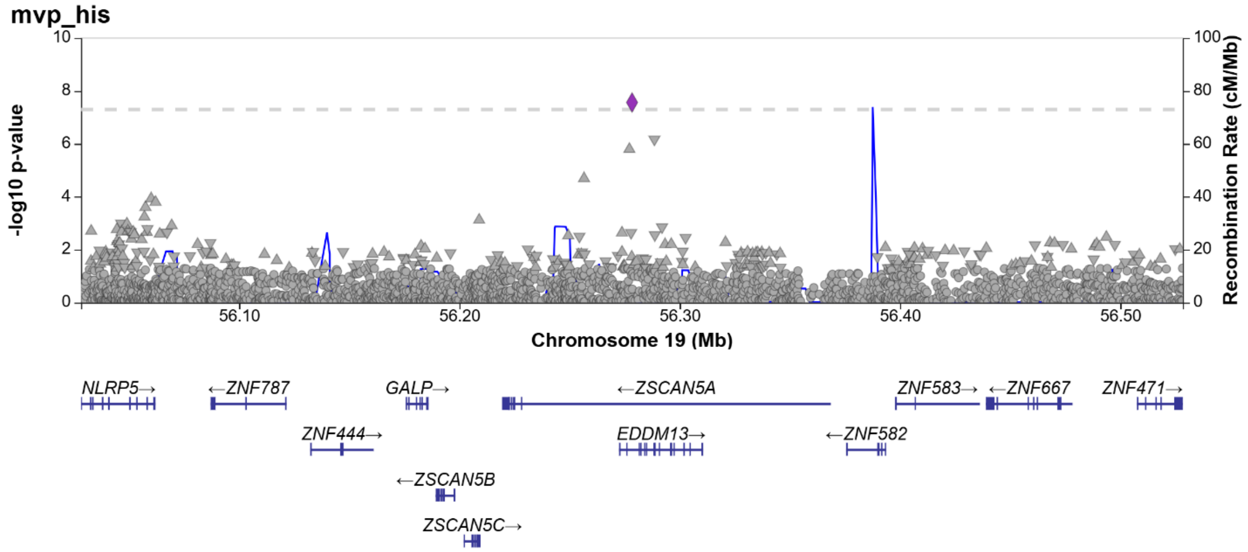

Figure S25. MAP3K1 in cross-ancestry

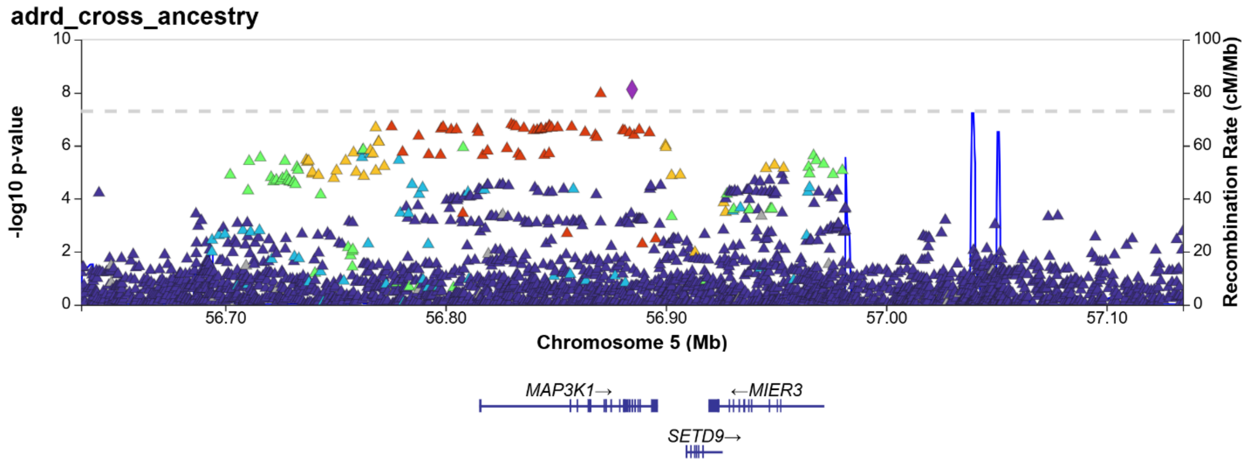


Figure S26. PTPRD in cross-ancestry

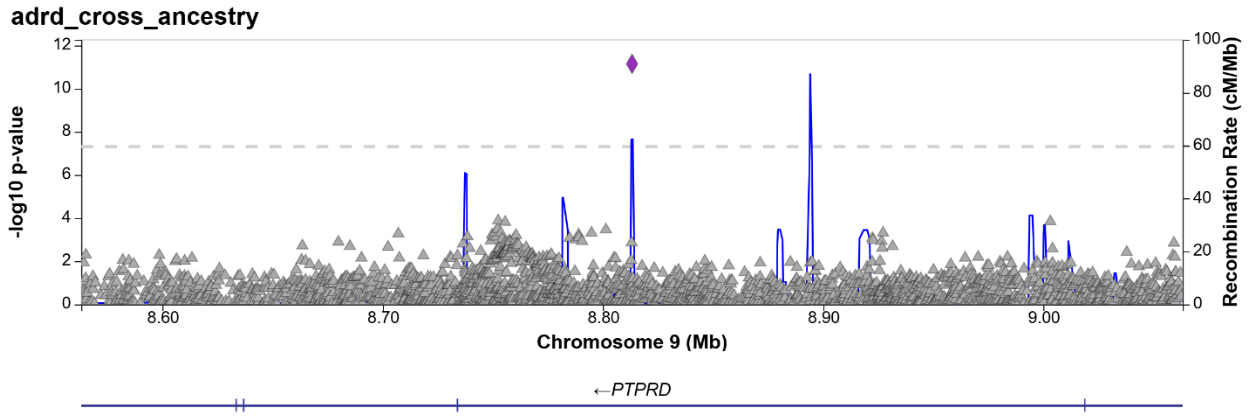

Figure S27. TRANK1 in cross-ancestry

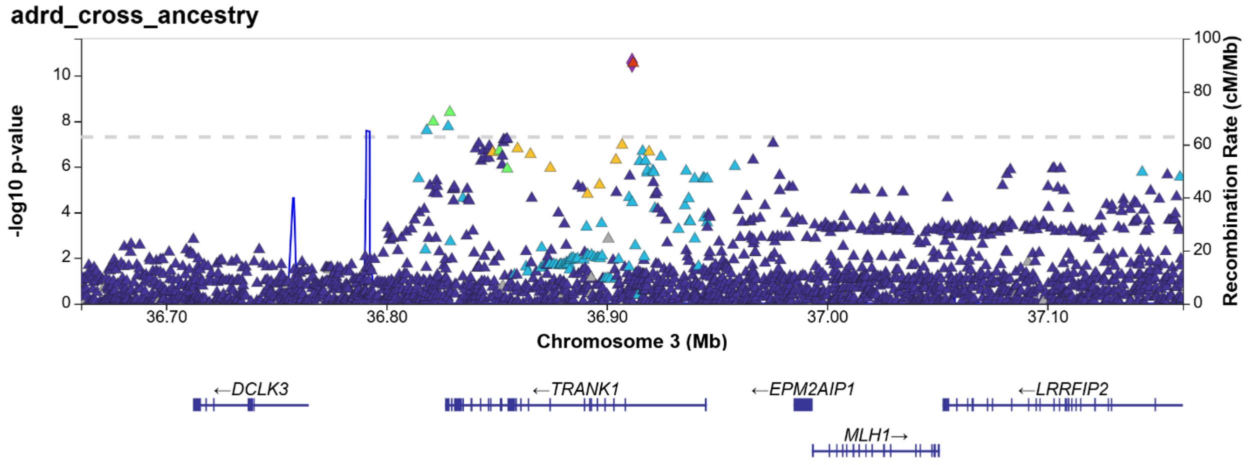


Figure S28. CAMK2D in cross-ancestry

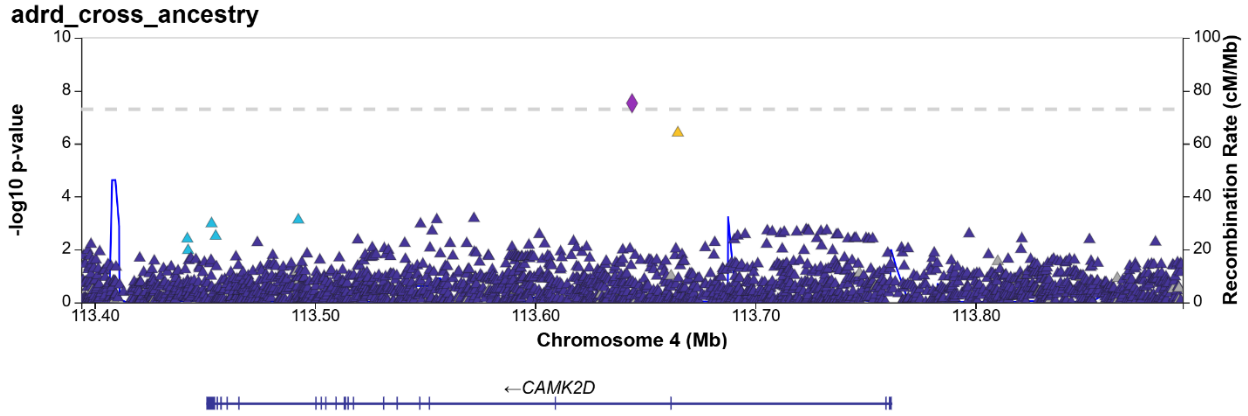

Figure S29. ZNF143 in cross-ancestry

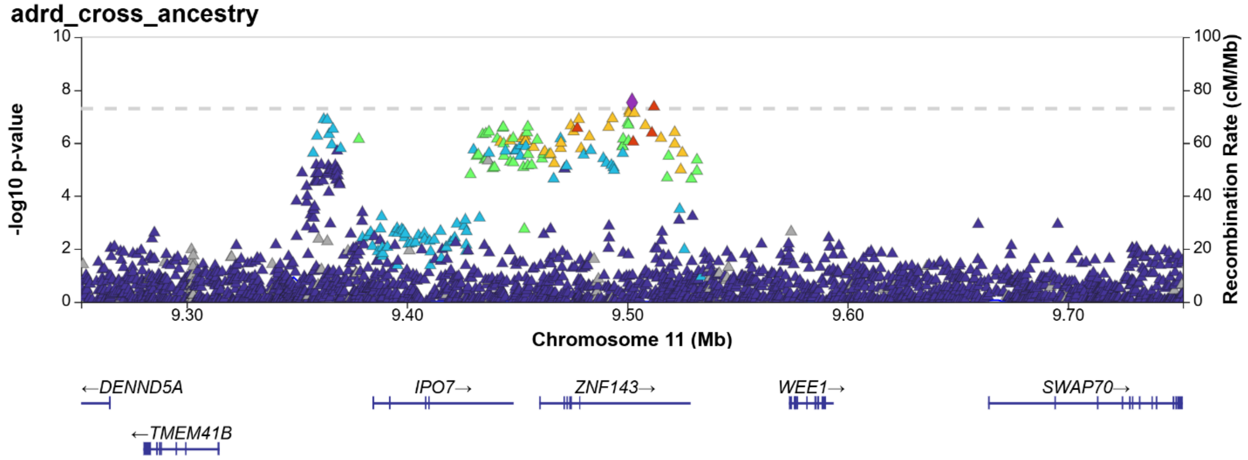


Figure S30. PRSS23 in cross-ancestry

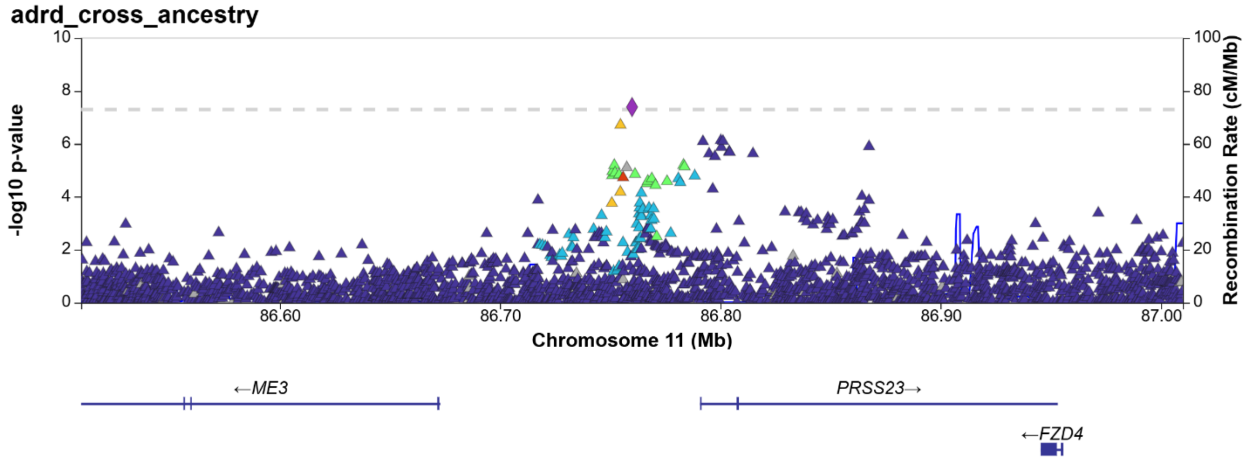

Figure S31. DLEU7 in cross-ancestry

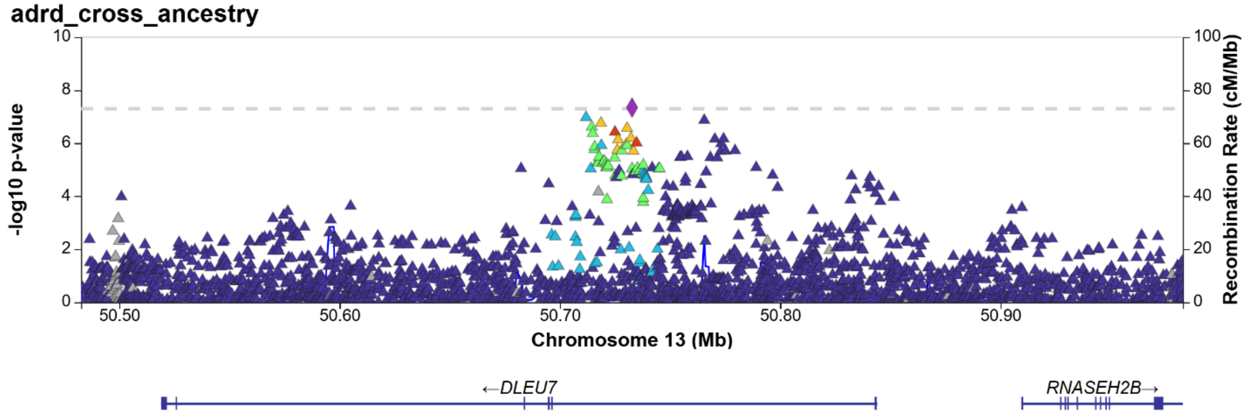


Figure S32. CCDC198 in cross-ancestry

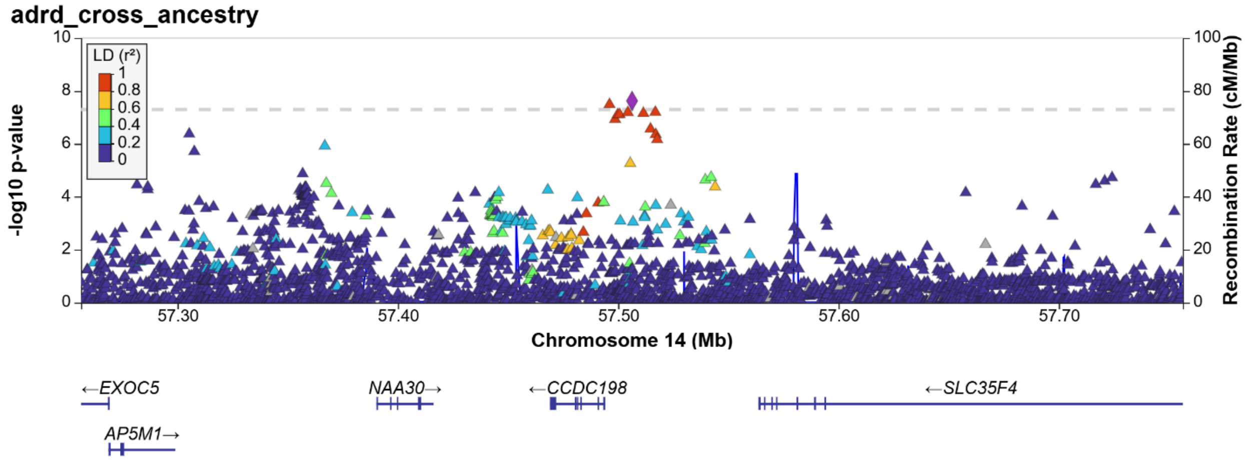

Figure S33. PPME1 in cross-ancestry

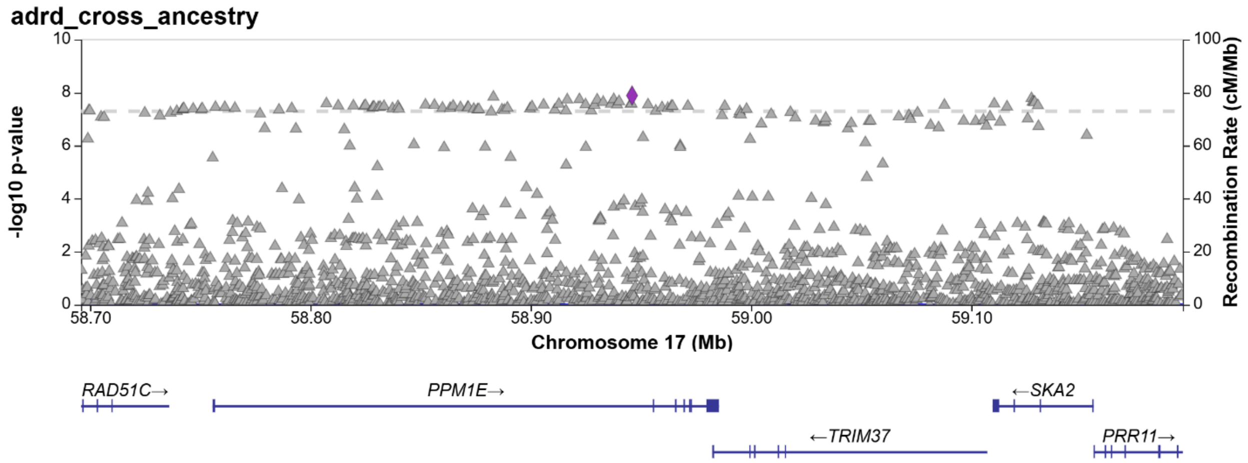


Figure S34. MYADAML2 in cross-ancestry

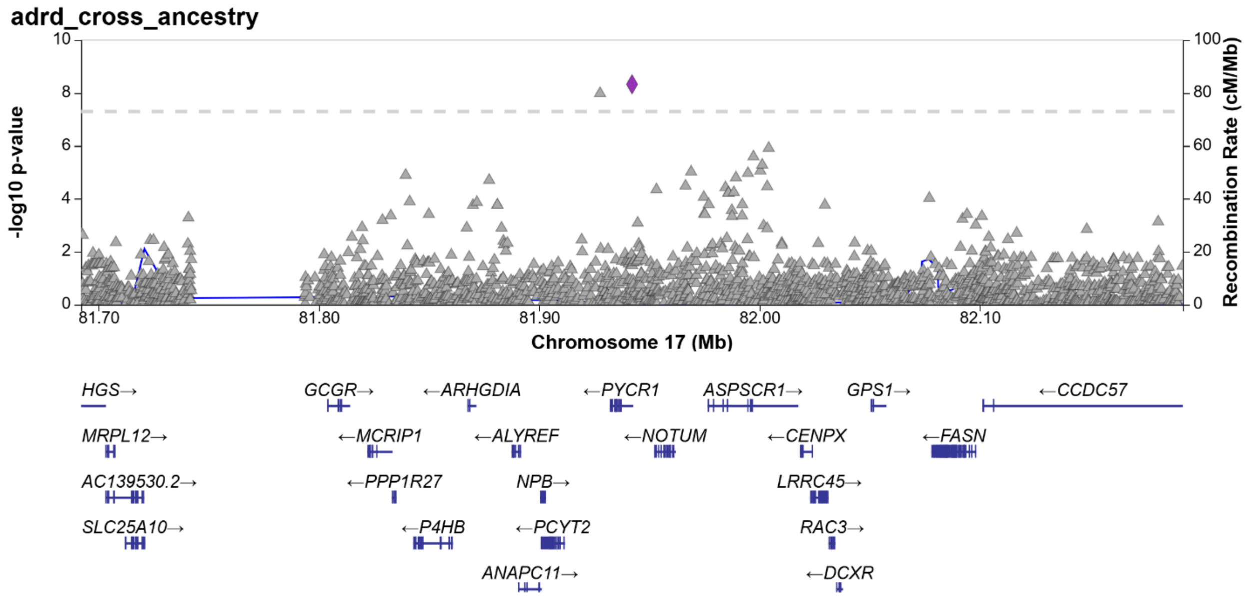

Figure S35. PLAUR in cross-ancestry

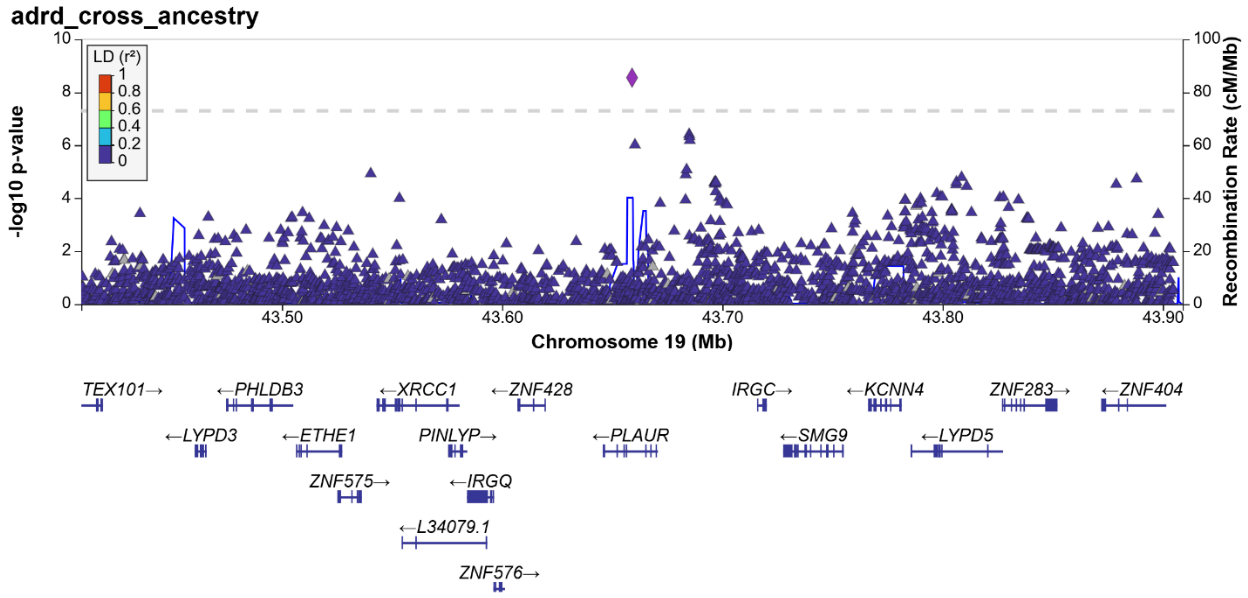


Figure S36. PAX7 in cross-ancestry

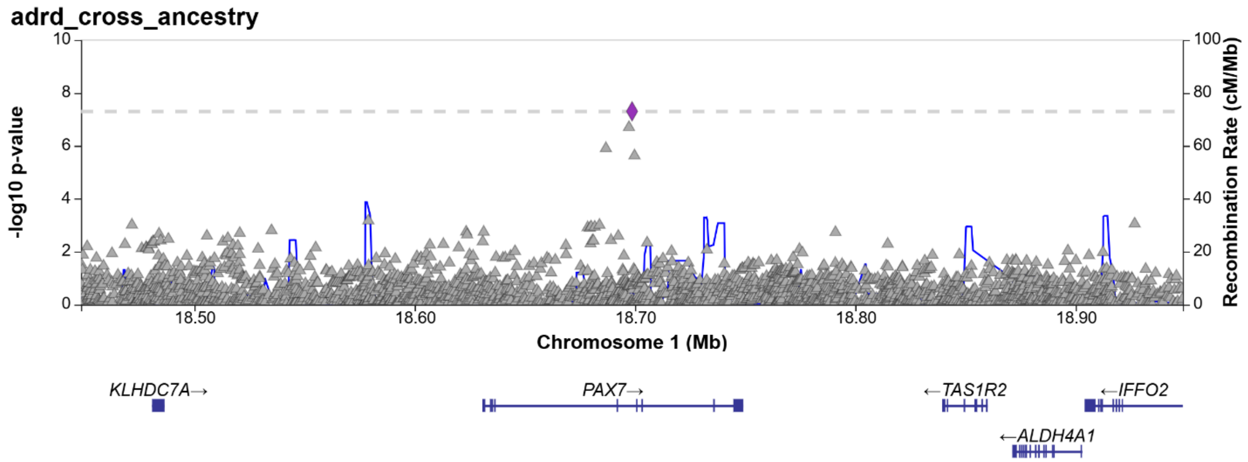

Figure S37. Garfield enrichment analysis results in the EA meta-analysis

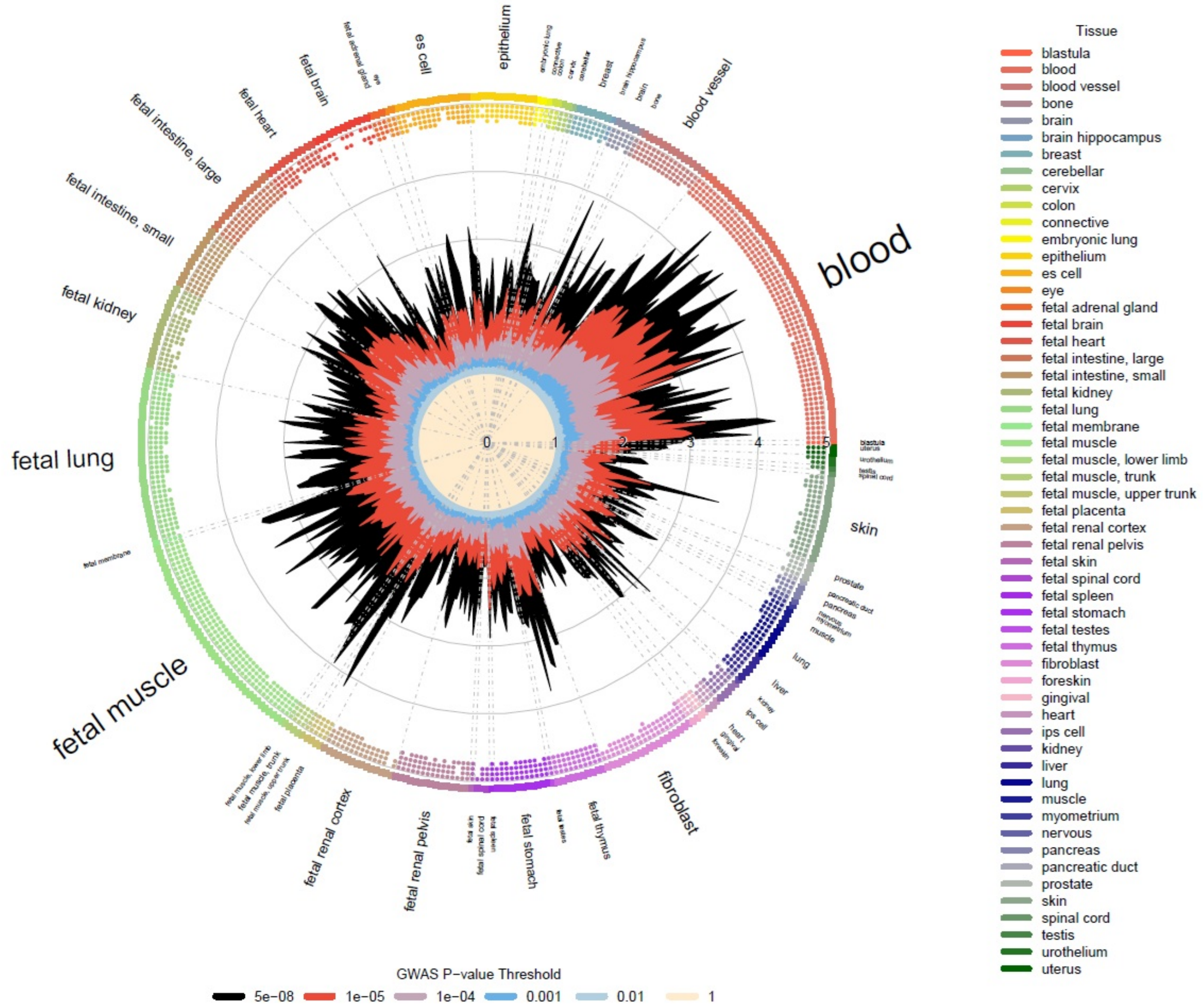


Enrichment of GWAS p-values in DNaseI hypersensitive sites. Radial lines show odds ratio values at six p-value thresholds (T) for all ENCODE and Roadmap Epigenomics DHS cell lines, sorted by tissue on the outer circle. Dots in the inner ring of the outer circle denote significant GARFIELD enrichment at the specified T value after multiple testing correction for the number of effective annotations and are colored with respect to the tissue of the cell type that they test. Font size of tissue labels reflects the number of cell types from that tissue

Figure S38: Results from the eQTL/GWAS hits colocalization analysis

Genome wide-significant SNPs are shown

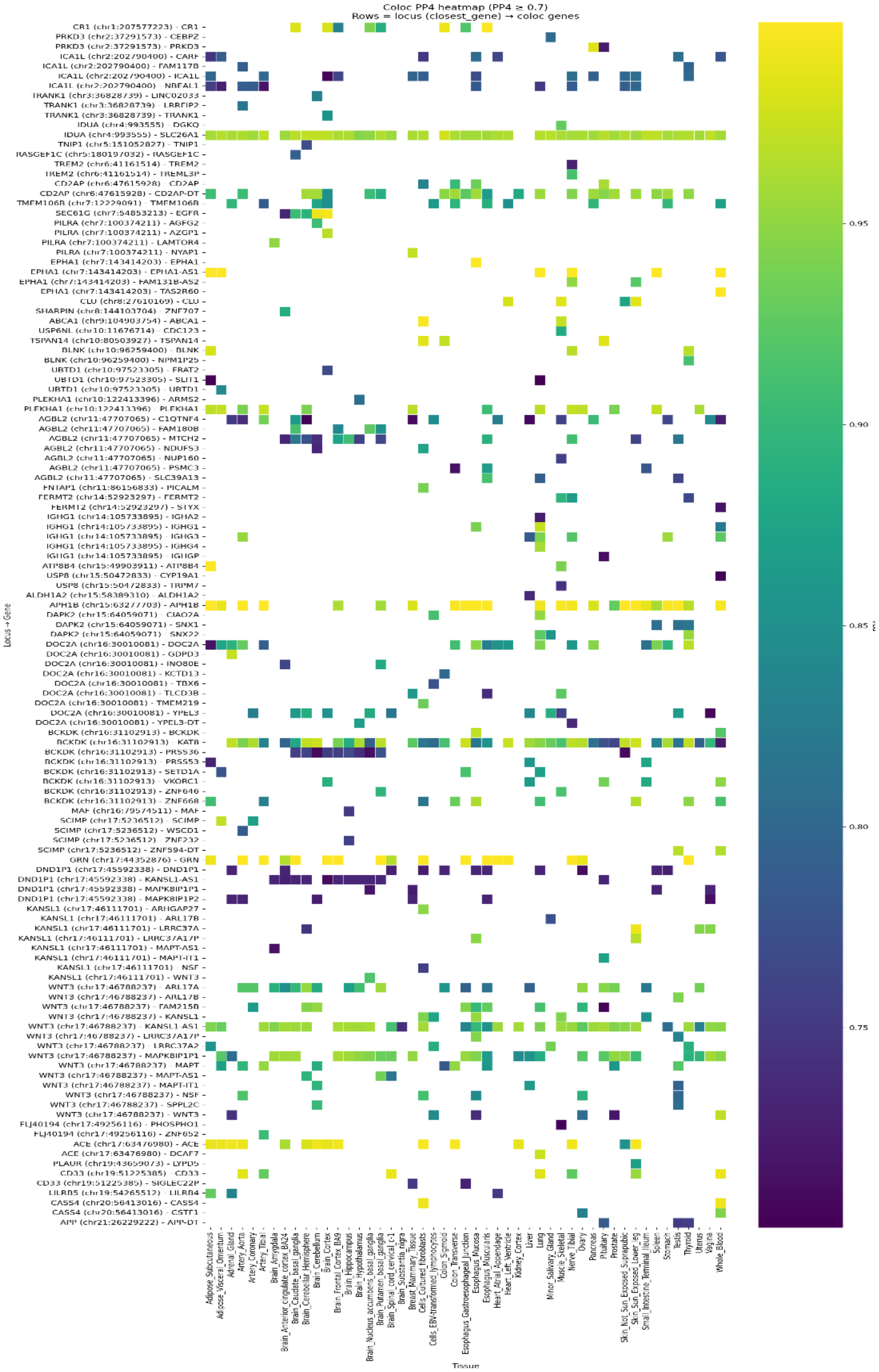

Genome wide-significant SNPs are shown on the left Y-axis. Tissues where those genes are expressed are on the X-axis. The colors of the squares indicate the significance of the evidence for co-localization according to the Bayes factors (right of Y-axis).
